# Redeployment Experiences of Healthcare Workers in the UK during COVID-19: data from the nationwide UK-REACH study

**DOI:** 10.1101/2024.03.03.24303615

**Authors:** Zainab Zuzer Lal, Christopher A. Martin, Mayuri Gogoi, Irtiza Qureshi, Luke Bryant, Padmasayee Papineni, Susie Lagrata, Laura B Nellums, Amani Al-Oraibi, Jonathon Chaloner, Katherine Woolf, Manish Pareek

## Abstract

**Background:** Increasing demands of COVID-19 on the healthcare system necessitated redeployment of HCWs outside their routine specialties. Previous studies, highlighting ethnic and occupational inequalities in redeployment, are limited by small cohorts with limited ethnic diversity.

**Aims:** To assess how ethnicity, migration status, and occupation are associated with HCWs’ redeployment experiences during COVID-19 in a nationwide ethnically diverse sample.

**Methods:** We conducted a cross-sectional analysis using data from the nationwide United Kingdom Research Study into Ethnicity And COVID-19 outcomes in Healthcare workers (UK-REACH) cohort study. We used logistic regression to examine associations of ethnicity, migration status, and occupation with redeployment experiences of HCWs, including provision of training and supervision, patient contact during redeployment and interaction with COVID-19 patients.

**Results:** Of the 10,889 HCWs included, 20.4% reported being redeployed during the first UK national lockdown in March 2020. Those in nursing roles (Odds Ratio (OR) 1.22, 95% Confidence Interval (CI) 1.04 – 1.42, p=0.009) (compared to medical roles) had higher likelihood of being redeployed as did migrants compared to those born in the UK (OR 1.26, 95% CI 1.06 - 1.49, p=0.01) (in a subcohort of HCWs on the agenda for change (AfC) pay scales). Asian HCWs were less likely to report receiving training (OR 0.66, 95% CI 0.50 – 0.88, p=0.005) and Black HCWs (OR 2.02, 95% CI 1.14 – 3.57, p=0.02) were more likely to report receiving supervision, compared to White colleagues. Finally, redeployed Black (OR 1.33, 95% CI 1.07 – 1.66, p=0.009) and Asian HCWs (OR 1.30, 95% CI 1.14 – 1.48, p<0.001) were more likely to report face-to-face interaction with COVID-19 patients than White HCWs.

**Conclusions:** Our findings highlight disparities in HCWs’ redeployment experiences by ethnicity, migration, and job role which are potentially related to structural inequities in healthcare. For future emergencies, redeployment should be contingent upon risk assessments, accompanied by training and supervision tailored to individual HCWs’ experience and skillset.

**What is already known on this topic:**
Ethnic minority healthcare workers (HCWs) were at an elevated risk of infection during COVID-19 due to occupational and socio-demographic factors. The strain on healthcare systems during the pandemic resulted in acute staffing shortages, prompting redeployment of HCWs to areas outside their professional training. However, recent research suggests inconsistent implementation of redeployment across ethnic groups, revealing structural disparities within the healthcare system.
**What this study adds:**
Our study, the largest of its kind, found no ethnic differences in the process of redeployment itself, but disparities emerged in the experiences of redeployment. Asian HCWs reported less likelihood of receiving training, while Black HCWs reported more likelihood of receiving supervision compared to their White counterparts. Ethnic minority HCWs were also more likely to report interaction with COVID-19 patients than their White colleagues. While there were no ethnic differences in the process of redeployment, occupational and migration differences reveal that those in nursing and midwifery roles (in comparison to medical roles), as well as migrant HCWs on the AfC payscale (in comparison to those born in the UK), were more likely to report being redeployed.
**How this study might affect research, practice or policy:**
This UK-wide study highlights inconsistencies in the redeployment process, training, supervision, and patient interactions based on occupation, ethnicity and migration status. Further investigation, incorporating qualitative and human resources data, is crucial to understand the complexities and address potential structural discrimination within the NHS. For future practice, redeployment should align with risk assessments and include training and supervision tailored to HCWs’ experience and skillset.

**Teaser text:** This study explores how ethnicity, migration status, and occupation were associated with healthcare workers’ (HCWs) redeployment experiences during COVID-19. After adjustment of covariates, we found that nursing roles and migration to the UK increase redeployment likelihood. Asian HCWs reported lesser training and Black HCWs reported more supervision, compared to White colleagues. Redeployed Black and Asian HCWs were more likely to report interaction with COVID-19 patients. Findings highlight disparities in HCWs’ redeployment experiences in an ethnically diverse sample.

## Introduction

The COVID-19 pandemic has caused over 6 million deaths worldwide as of August 2023 [1]. People from ethnic minority backgrounds have had a significantly higher risk of contracting COVID-19 and dying from it [2]. In the UK, COVID-19 mortality rates have been higher in Bangladeshi, Pakistani, and Black Caribbean communities compared to the White British group [3]. Front-line healthcare workers (HCWs) have also faced a disproportionate risk of COVID-19 compared to the general population, with a previous study finding a threefold higher risk of SARS-CoV-2 infection which may have been exacerbated by the inadequate availability of personal protective equipment (PPE) [4–7]. The intersection of heightened risk and increased rates of infection in ethnic minority groups may be mediated by occupational and sociodemographic factors, which, in turn, are manifestations of structural discrimination [8,9]. These factors include inhabiting densely populated areas, living in multigenerational households, and occupying housing with poorer ventilation [10,11]. Therefore, within ethnic minority healthcare workers, the convergence of ethnicity and occupation puts them at higher risk of COVID-19 infection, morbidity, and mortality [8,12]. The impact of the pandemic on healthcare systems has also been profound, with acute staffing shortages, limited number of beds, growing waitlists and increased caseloads [13]. In response to these escalating demands, various strategies were implemented in April 2020, including staff mobilisation, redeployment to areas outside their professional training, alteration of work schedules, and risk assessments to assess individual risk factors for COVID-19 and concerns of employees [14,15]. Redeployment, specifically, was employed for reassigning healthcare workers to alternative units or speciality areas [16]. Although redeployment played a critical role in effectively managing the crisis, recent research suggests it significantly affected staff well-being, with over 95% HCWs reporting stress and anxiety after being redeployed [17]. Previous studies have highlighted ethnic inequalities in redeployment, with HCWs from ethnic minority backgrounds being more likely to be redeployed to COVID-19 areas than their White counterparts [17,18]. However, these studies were conducted in small cohorts with a low proportion of participants from ethnic minority groups and did not examine outcomes relating to experiences of redeployment such as the degree of patient contact in the redeployed role or the receipt of training/supervision [17,18].

Therefore, to address this knowledge gap, we conducted an analysis using data from a nationwide cohort study of UK HCWs, the UK-REACH Study (UK Research study into Ethnicity And COVID-19 outcomes in Healthcare workers). Our aims were to determine whether ethnicity, migration status or occupational role affected the likelihood of redeployment. Amongst those redeployed, we aimed to examine the receipt of training and supervision before/during redeployment, the level of direct patient contact in the redeployed role in comparison to the usual role, and whether the role involved COVID-19 contact.

## Methods

UK-REACH, a nationwide cohort study comprises multiple work packages to understand the impact of COVID-19 on HCWs from diverse ethnic backgrounds. Here, we used data from the baseline questionnaire of the study.

### Recruitment

We included HCWs (including ancillary workers in healthcare settings) aged 16 and above. The first stage of recruitment involved healthcare regulators (see supplementary information for a list of participating regulators) sending email invitations to their registrants. We directed interested HCWs to the UK-REACH website (https://www.uk-reach.org) where they could access the participant information sheet and provide informed consent. We then asked participants to complete the online questionnaire. We supplemented this sample by direct recruitment from participating NHS Trusts (see study protocol and cohort profile for further details [19,20]).

We administered the baseline questionnaire between 4th December 2020 and 8^th^ March 2021.

### Defining the analysed cohort

Formation of the analysed sample is shown in Figure 1.

**Figure 1:**
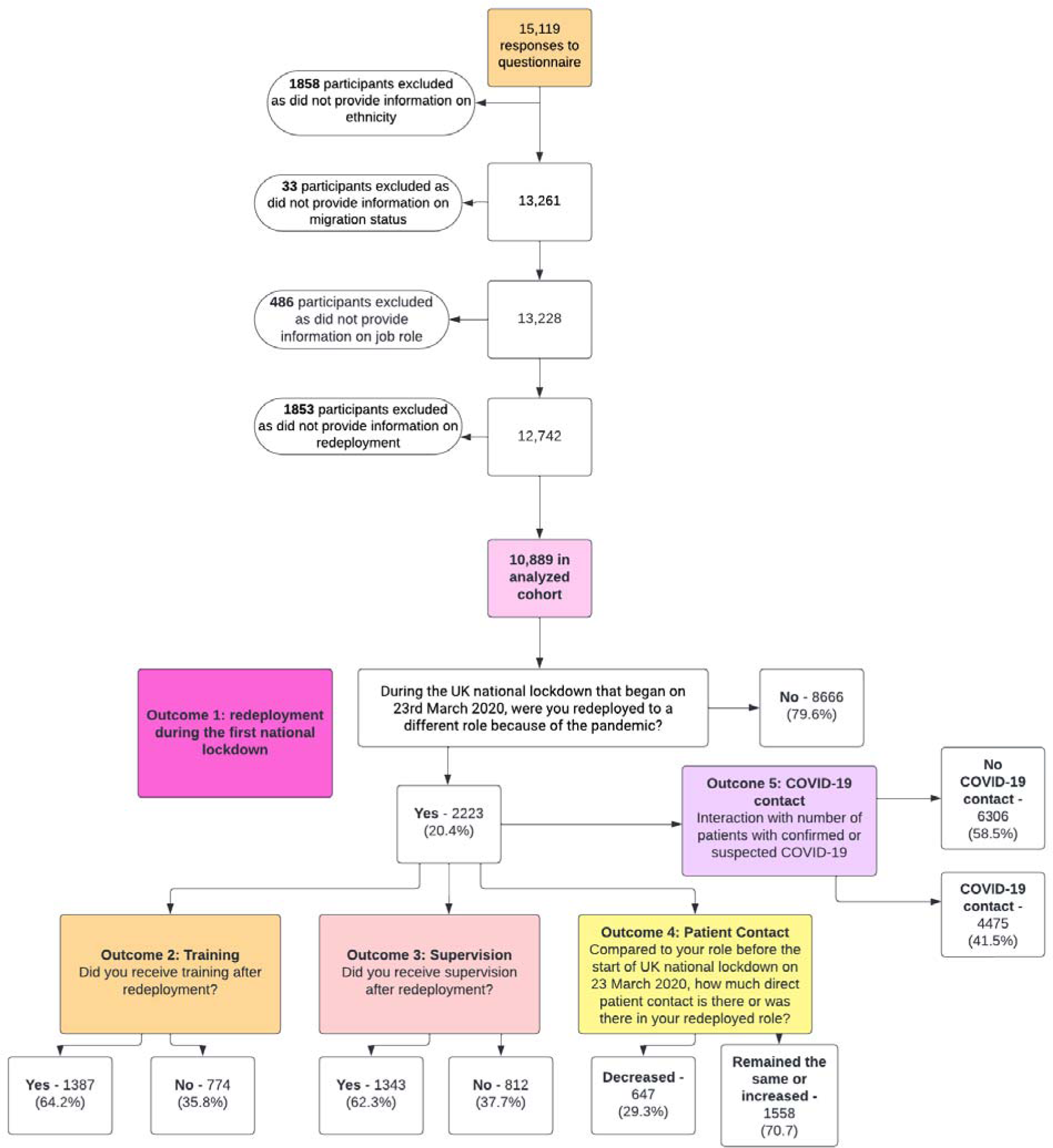
Formation of the analysed sample and derivation of outcome measures.

We excluded participants who did not provide information on ethnicity, migration status, occupation, and redeployment. We asked participants about redeployment experiences during the first UK national lockdown and therefore excluded anyone who indicated they were not working during this period. To ensure our measures of COVID-19 work patterns reflected experiences at the time of staff deployment, we used answers to questions about occupational circumstances in the weeks following the UK national lockdown (referred to as “the first UK national lockdown on 23 March 2020”, throughout the questionnaire).

### Outcome measures

We derived 5 binary outcome measures from questionnaire items (Figure 1). Outcome 1 is derived from the question ‘During the UK national lockdown that began on 23rd March 2020, were you redeployed to a different role because of the pandemic?’. Outcomes 2 & 3 are derived from the questions ‘Did you receive training during redeployment?’ and ‘Did you receive supervision during redeployment?’. Outcome 4 is derived from the question ‘Compared to your role before the start of UK national lockdown on 23 March 2020, how much direct patient contact is there or was there in your redeployed role?’. Outcome 5 is not derived from a question that directly relates to redeployment, but rather a questionnaire item asking HCWs to declare the number of face-to-face interactions with COVID-19 patients in a typical week during the first month after the start of the lockdown (see Figure 1 for further details). Both, redeployment and the COVID-19 patient contact items ask about the same period and it is assumed therefore that HCWs who were redeployed provided information on COVID-19 patient contact in their redeployed roles.

Outcome 1: redeployment (redeployed vs not redeployed);

Outcome 2: received training during redeployment (received vs did not receive); Outcome 3: received supervision during redeployment (received vs did not receive);

Outcome 4: change in patient contact during redeployment (stayed the same or decreased vs increased);

Outcome 5: interaction with number of patients with confirmed or suspected COVID-19 (no COVID-19 patient contact vs COVID-19 patient contact).

For analysis of outcomes 2, 3, 4, and 5, we only included those who answered they were redeployed.

### Exposures

Our exposures of interest were self-reported ethnicity, migration status, and occupation.

**- Ethnicity:** categorised into five broad ethnic groups, as suggested by the UK Office for National Statistics (White, Asian, Black, Mixed, and Other) [21], to increase statistical power.
**- Migration status**: whether participants were born in the UK or outside the UK.
**- Occupation**: categorised as doctors, nursing staff (including midwives and nursing associates), allied health professionals, pharmacy staff, healthcare scientists, ambulance staff, dental, optical, administrative, estates, facilities or other wider healthcare roles, and others (including medical associates).

### Covariates

Based on existing literature and expert opinion, potential confounders of the relationship between our exposure and outcome measures were hypothesized to be:

**- Demographic characteristics** (age categorised into 16-30-year-olds, 31-45, 46-60, and 61 and above; sex categorised into male and female),
**- Occupational factors** (job sector – i.e., worked for the NHS in any capacity, or worked outside the NHS)
**- Deprivation** – measured by the index of multiple deprivation [IMD, the official measure of relative deprivation for small areas or neighbourhoods in England, expressed as quintiles] [22]
**- Self-reported long-term health conditions** (LTCs) categorised as those requiring shielding – including organ transplant, diabetes, asthma, heart disease, kidney disease, liver disease, cancer, and immunosuppression (based on the COVID-19 guidance for people who were considered at higher risk)[23], other conditions (including hypertension, obesity, stroke, conditions affecting the brain or nervous system, and mental health conditions), and no conditions.

### Subgroup analyses

To control for the effect of more granular occupational variables and occupational seniority we conducted analyses on two subgroups; doctors and those on the NHS agenda for change (AfC) pay scales. The NHS AfC pay bands comprise bands 1-9 (with salary increasing as band level rises) for HCWs other than doctors, dentists and very senior managers. This scale was used as a proxy measure for occupational seniority. We repeated analyses of outcomes 1, 2 and 3 within these subgroups adjusting for the grade or stage of training for doctors and the AfC pay band for those on these pay scales.

### Statistical analysis

All variables were categorical were summarised as frequency and percentage. The derivation of all variables used in the analysis is highlighted in Supplementary Table 1.

We used univariable and multivariable logistic regression models to explore relationships between the exposure and outcome variables. The results are presented in terms of odds ratios (ORs) and adjusted odds ratios (aORs), 95% confidence intervals (CI), and p-values. Adjusted models included ethnicity, occupation, and migration status, along with confounders (age, sex, LTCs, and deprivation). In subgroup analyses of specific occupational groups, we added AfC pay band (for those in non-medical roles) and grade (for doctors). For analysis of outcomes 2 & 3, we did not include the underlying comorbidities variable since we believed they would not impact whether healthcare workers received training and supervision during redeployment.

All analyses were conducted using Stata V.17.

### Missing data

We presented the frequency and proportion of missing data for each variable used in the analysis. We used multiple imputation by chained equations to impute missing data in the logistic regression models. In the imputation models, we included all variables used in the analysis of outcome 1 (including the outcome measure), except the one being imputed. We applied Rubin’s Rules to combine parameter estimates and standard errors from 10 imputations into a single set of results [24]). We used a random number seed to ensure reproducible results. In subgroup analyses including grade or pay band, we excluded those who did not provide this information.

### Ethics

The study was approved by the Health Research Authority (Brighton and Sussex Research Ethics Committee; ethics reference: 20/HRA/4718). All participants gave written informed consent.

### Involvement and engagement

A Professional Expert Panel of HCWs from a diverse range of ethnic backgrounds, healthcare roles, and genders, both locally and nationally, worked closely to help develop the research question, analysis plan and manuscript.

## Results

### Recruitment and formation of analysis sample

Figure 1 shows the formation of the analysed sample. In total, 15,119 HCWs responded to the questionnaire. After excluding 4230 participants due to missing data on ethnicity, migration status, job role, and redeployment (as detailed in Figure 1) – we arrived at our final analysis sample of 10,889 HCWs.

### Description of the analysed cohort

Table 1 summarises the analysed cohort in terms of demographic, household, and occupational factors together with the amount of missing data for each variable. The majority of those included were women (75.2%) and 30.0% were from ethnic minority groups (19.0% Asian, 4.2% Black, 4.1% Mixed, 2.0% Other). Approximately 25% were doctors, 23% worked in nursing and midwifery roles, and 30% in allied health professional roles.

**Table 1.** Description of the analysed cohort. Table 1 provides a description of the 10,889 HCWs who worked during the first UK national lockdown starting 23^rd^ March 2020. These HCWs provided information on their ethnicity, migration status, job role, and answered questions about redeployment. All data in the right-hand column are n (%). *Include organ transplant, diabetes, asthma, heart disease, kidney disease, liver disease, cancer, and immunosuppression (based on the COVID-19 guidance^18^ for people who were considered at higher risk). HCWs, healthcare workers; NHS, National Health Service

| Variable: | Description:<br>N = 10,889 |
| --- | --- |
| **Demographic and household factors** |  |
| **Ethnicity** |  |
| White | 7683 (70.6) |
| Black | 460 (4.2) |
| Asian | 2076 (19) |
| Mixed | 448 (4.1) |
| Other | 222 (2) |
| **Age** |  |
| <30 years | 1596 (14.7) |
| 31-45 years | 3987 (36.1) |
| 46-60 years | 4319 (39.7) |
| 61 and above | 933 (9) |
| Missing | 54 (0.5) |
| **Sex** |  |
| Male | 2676 (24.6) |
| Female | 8187 (75.2) |
| Missing | 26 (0.2) |
| **Migration status** |  |
| Born in the UK | 8000 (73.5) |
| Born overseas | 2889 (26.5) |
| **Index of Multiple Deprivation (quintiles)** |  |
| 1 (most deprived) | 951 (9.9) |
| 2 | 1624 (16.9) |
| 3 | 1971 (20.5) |
| 4 | 2339 (24.3) |
| 5 (least deprived) | 2740 (28.5) |
| Missing | 1264 (11.6) |
| **Underlying health conditions** |  |
| No health conditions | 4573 (42) |
| Conditions that required shielding during COVID-19* | 2107 (19.4) |
| Other conditions | 3232 (29.7) |
| Missing | 977 (9.0) |
| **Occupational factors** |  |
| **Occupation** |  |
| Doctors | 2688 (24.7) |
| Nursing | 2485 (22.8) |
| Allied Health Professionals | 3307 (30.4) |
| Pharmacy | 233 (2.1) |
| Healthcare scientist | 532 (4.9) |
| Ambulance | 442 (4.1) |
| Dental | 435 (4) |
| Optical | 117 (1.1) |
| Administrative | 622 (5.7) |
| Estates/ facilities | 103 (1) |
| Other | 320 (3) |
| **Job sector:** |  |
| Worked for the NHS in any capacity | 9056 (83.2) |
| Worked outside the NHS | 1387 (12.7) |
| Missing | 446 (4.1) |
| **Redeployment experiences** |  |
| **Redeployment** |  |
| Redeployed | 2223 (20.4) |
| Not redeployed | 8666 (79.6) |
| **Training redeployed staff:** |  |
| Received training | 1387 (64.2) |
| Did not receive training | 774 (35.8) |
| **Supervising redeployed staff:** |  |
| Received supervision | 1343 (62.3) |
| Did not receive supervision | 812 (37.7) |
| **Patient contact during redeployment:** |  |
| Decreased | 647 (29.3) |
| Remained the same or increased | 1558 (70.7) |
| **COVID-19 contact** |  |
| No COVID-19 contact | 6306 (58.5) |
| COVID-19 contact | 4475 (41.5) |

### Univariable analysis of predictor variables

Univariable analysis of redeployment is shown in Supplementary Table 3, while Supplementary Table 4 presents the analysis in a sub-cohort of doctors and HCWs on the AfC pay scale to control for seniority. Overall, 2223 (20.4%) of the 10,889 HCWs who worked during the lockdown reported being redeployed. HCWs from Mixed ethnic groups, as compared to those from White groups had higher odds of reporting redeployment (OR 1.28, 95% CI 1.03 – 1.60, p=0.03). HCWs working in pharmacy (0.41, 0.27 – 0.63, p<0.001), healthcare scientist (0.43, 0.32 – 0.57, p<0.001), ambulance (0.5, 0.37 – 0.67, p<0.001), dental (0.75, 0.57 – 0.97, p=0.03), optical (0.53, 0.30 – 0.91, p=0.02) and administrative (0.46, 0.30 – 0.70, p<0.001) roles were less likely to report redeployment compared to doctors.

### Multivariable analysis

#### Redeployment

Ethnic differences seen in the univariable model attenuated after adjustment for covariates (Figure 2 and Supplementary Table 5). Compared to doctors, those working in nursing and midwifery (1.22, 1.04 – 1.42, p=0.009), and allied health professional roles (1.23, 1.07 – 1.41, p=0.003) were more likely to report being redeployed.

**Figure 2.**
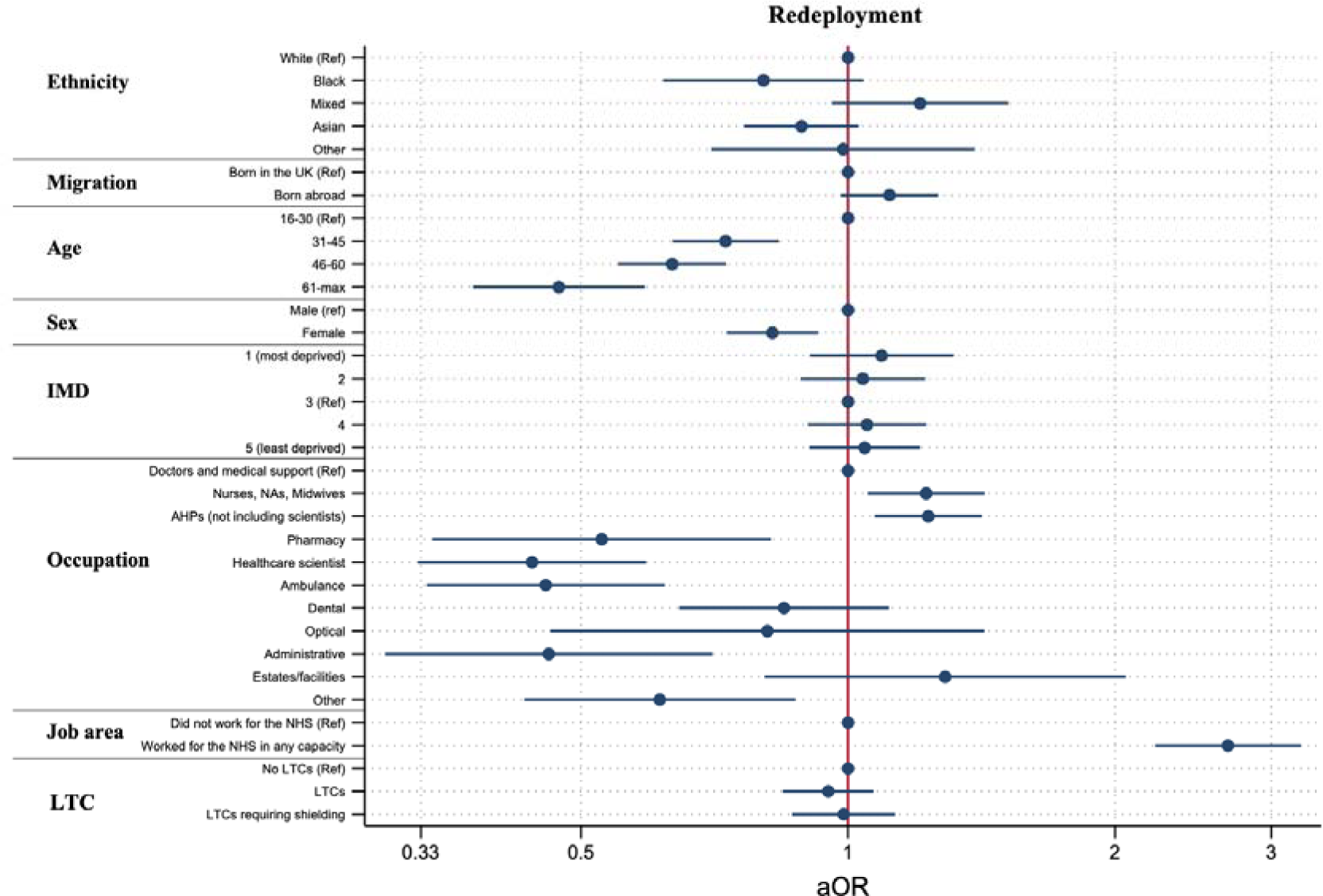
The relationship between ethnicity, migration status, and occupation with redeployment after adjustment for demographic and health covariates.

#### Training and supervision in redeployed roles

Of the 2223 HCWs who reported being redeployed, 62 did not respond to follow-up questions about training and 68 did not respond to questions about supervision. Among the redeployed HCWs, 64.2% (n=1387) reported receiving training while 62.3% (n=1343) reported receiving supervision. Asian HCWs were less likely to report training during redeployment compared to White HCWs (0.66, 0.50 – 0.88, p=0.005). HCWs born in the UK were more likely to report receiving training than those born in the UK (1.30, 1.01 – 1.66, p=0.04) (Figure 3 and Supplementary Table 5).

**Figure 3.**
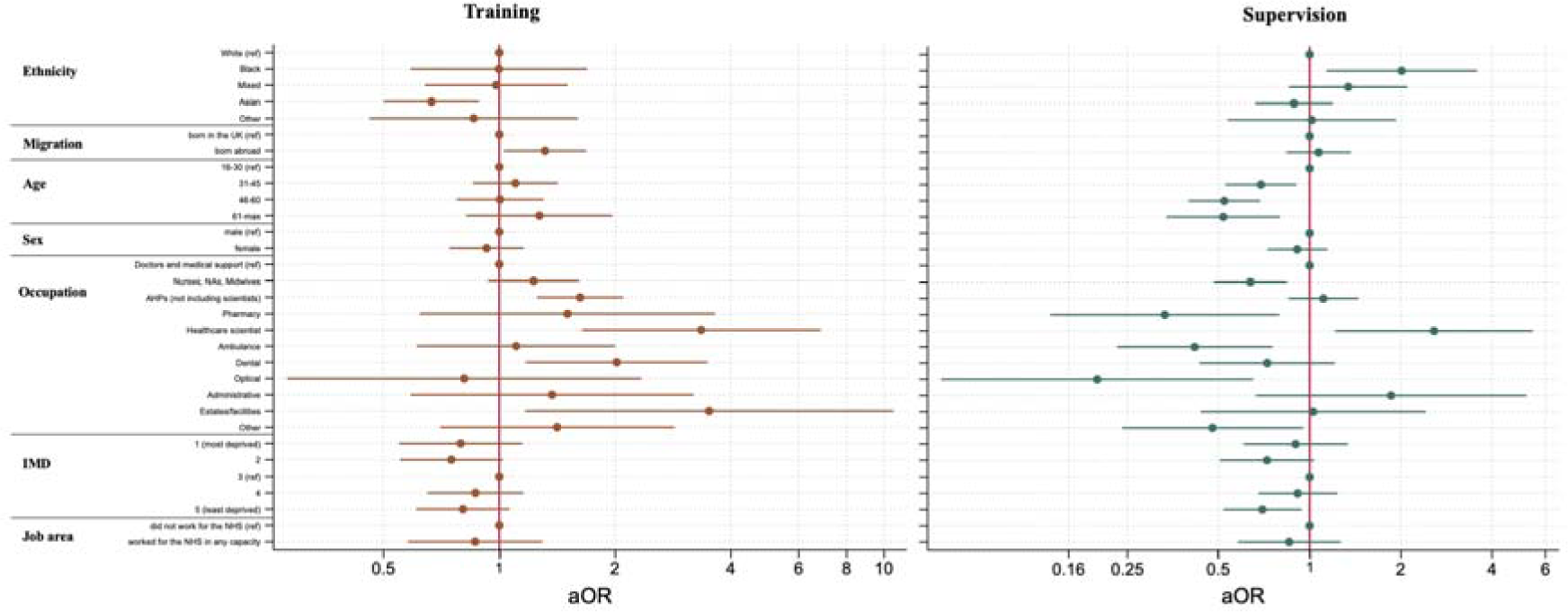
The relationship between ethnicity, migration status, and occupation with training and supervision after adjustment for demographic and health covariates.

Black HCWs were significantly more likely to report being supervised in their redeployed role (2.02, 1.14 – 3.57, p=0.02) compared to their White counterparts. Additionally, those living in the least deprived areas were less likely to report supervision than those from most deprived (IMD quintile 5: 0.71, 0.54 – 0.95, p=0.02 vs quintile 1) (Figure 3 and Supplementary Table 5).

#### Patient contact during redeployment

Nursing and midwifery roles, and allied health professional roles, in comparison to doctors, reported increased patient contact during redeployment (1.52, 1.1 – 2.08 p=0.01), whereas most other HCW groups reported decreased patient contact in their redeployed roles. HCWs with LTCs that required them to shield during the pandemic had less patient contact during redeployment than those without such conditions (0.62, 0.48 – 0.79, p<0.001) (Figure 4 and Supplementary Table 5).

**Figure 4.**
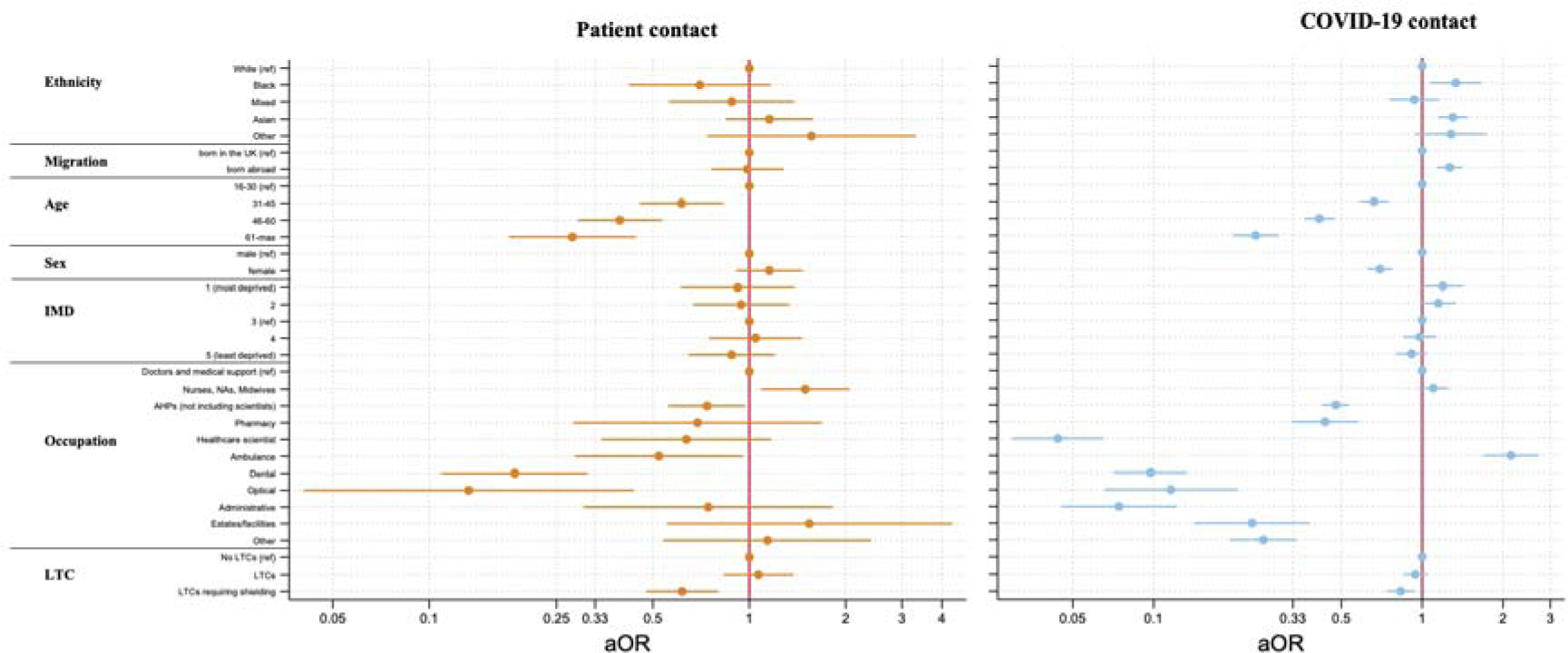
The relationship between ethnicity, migration status, and occupation with patient contact and COVID-19 contact after adjustment for demographic and health covariates. **Figures 2, 3 and 4** show adjusted ORs and 95% CIs, derived from multivariable logistic regression models, for the association of socio-demographic and occupational variables with 5 outcome measures relating to redeployment and redeployment experiences. ORs are adjusted for age, sex, ethnicity, occupation, migration, deprivation, and underlying long-term health conditions. All variables only include HCWs who were working during the UK national lockdown that began on 23^rd^ March 2020. The analysed sample included all HCWs who provided information about their ethnicity, migration status, job role, and whether or not they were redeployed. Outcome 1, a binary variable, is whether or not a HCW was redeployed to a different role because of the pandemic. Outcome 2, a binary variable is whether or not a HCW received training during redeployment. Outcome 3, a binary variable, is whether or not a HCW received supervision during redeployment. Outcome 4, a binary variable is whether direct patient contact decreased, remained the same or increased in HCW’s redeployed role. Outcome 5, a binary variable, is whether or not a HCW interacted with patients with COVID-19. Derivation of all outcomes is described in detail in **figure 1**. In the occupation variable, nursing includes midwives and nursing associates; HCW, healthcare workers; NHS, National Health Service.

#### Interaction with COVID-19 patients among redeployed HCWs

Those from a Black (1.33, 1.07 – 1.66, p=0.009) and Asian (1.30, 1.14 – 1.48, p<0.001) ethnic background were significantly more likely to report interaction with COVID-19 patients than White HCWs (Figure 4 and Supplementary Table 5). Similarly, HCWs born outside the UK (1.27, 1.13 – 1.42, p<0.001), as opposed to those born in the UK were more likely to report increased interaction with COVID-19 patients.

#### Sub-cohort analyses

Supplementary table 2 describes the cohort based on their level of seniority using the AfC pay band and doctors’ grades as proxy measures for occupational seniority. About 19% of HCWs on the AfC pay scale worked in Band 6 (n=2029), and 9% of doctors held consultant positions (n=966).

##### Redeployment

Among those on the AfC pay scale, we found that HCWs in senior positions, like Band 8 or 9 (0.69, 0.55 - 0.87, p=0.001), in comparison to junior roles like Band 5, were less likely to report redeployment. Further, HCWs born in the UK were less likely to report redeployment than those born abroad (1.26, 1.06 – 1.49, p=0.01) (Supplementary Table 7 & Supplementary Figure 2).

After adjusting for doctor’s grade, consultants were significantly less likely to report being redeployed in comparison to foundation-year doctors (3.54, 2.15 - 5.83, p<0.001).

Additionally, doctors from a Black ethnic background were less likely to report redeployment (0.58, 0.37 – 0.91, p=0.02) compared to their White colleagues (Supplementary Table 6 & Supplementary Figure 1).

##### Training and supervision

Doctors of Asian background (0.55, 0.34 – 0.87, p=0.01) were less likely to report receiving training for their redeployed role, however, this effect was not observed in Asian HCWs within the AfC pay scale. Doctors from a Black background (5.41, 1.10 – 26.60, p=0.04) were significantly more likely to report receiving supervision compared to their White colleagues, which again was not evident within HCWs on the AfC pay scale (Supplementary Table 6 & Supplementary Figure 3).

##### COVID-19 contact

After adjusting for grade, the significant effect of ethnicity on COVID-19 patient contact diminished, however, doctors born overseas were more likely to report contact with COVID-19 patients (1.40, 1.14 – 1.72, p=0.001) (Supplementary Table 6 & Supplementary Figure 4). Among HCWs on the AfC pay scale, Black (1.62, 1.15 – 2.27, p=0.006) and Asian (1.24, 1.00 – 1.52, p=0.05) HCWs were significantly more likely to report COVID-19 contact. (Supplementary Table 7 & Supplementary Figure 5).

## Discussion

Our study, the largest of its kind, analyses data from a diverse sample of over 10,000 HCWs in the UK and sheds light on variations in redeployment experiences. We identified inconsistencies in the redeployment process, training, supervision, patient contact, and interaction with COVID-19 patients based on occupation, ethnicity, and migration status. Whilst we found no differences in likelihood of redeployment by ethnicity in the cohort overall, our results suggest migrant HCWs on AfC pay bands were more likely to be redeployed than those born in the UK. We found that in comparison to White HCWs, those from an Asian background were less likely to report receiving training during redeployment, while Black HCWs were more likely to receive supervision. Further, redeployed ethnic minority and migrant HCWs, were more likely to report contact with COVID-19 patients compared to those from a White ethnic background and born in the UK respectively.

As per existing literature [18,25] our findings indicate that occupation played a crucial role in the likelihood of being redeployed. Compared to those in medical roles, HCWs in nursing, midwifery, and allied health professional roles were more likely to report redeployment, while those in pharmacy, healthcare scientist, ambulance, dental, optical, and administrative roles were less likely. This discrepancy may be attributed to differences in skill sets acquired during training and the diverse career paths that HCWs follow after their qualification. Lower reports of redeployment among certain HCWs may be due to the potential to transition to remote work or the absence of direct patient care-related skills (as in the case of scientific staff) [26].

Our study highlights the challenges faced by HCWs in nursing roles, revealing increased patient contact in redeployed roles compared to doctors, potentially contributing to higher occupational exposure to SARS-CoV-2 and greater risk of infection [26]. Existing literature has already highlighted the adverse impact of the pandemic on their physical and mental well-being, noting increased susceptibility to COVID-19, and elevated stress and anxiety levels [27,28]. There are likely to be several factors underlying these observations including inadequate training, concerns about safety, shortages of personal protective equipment (PPE), and gaps in support and structured communication [28,29].

Our study highlights that HCWs from migrant backgrounds on the AfC pay band, were more likely to be redeployed compared to their counterparts born in the UK. Additionally, HCWs from an ethnic minority background were significantly more likely to report interaction with COVID-19 patients during the UK national lockdown close to the peak of the first wave. These findings raise serious concerns as HCWs from ethnic minority backgrounds already faced several compounding risk factors. They were at a higher risk of contracting COVID-19, and despite this increased risk, had limited access to risk assessments compared to HCWs from a White background, as revealed by the UK-REACH qualitative study [26,30]. It is crucial to note that our quantitative findings suggest that HCWs from an ethnic minority background were offered risk assessments but experienced no changes in their working practices [31]. This could be linked to structural discrimination as literature extensively documents that HCWs from ethnic minority backgrounds often felt less empowered to voice concerns about risk assessments, and had less autonomy in decisions related to their redeployment [25,32]. Furthermore, HCWs from an ethnic minority background are more likely to work in junior roles involving increased patient contact [9]. With evidence indicating insufficient access to personal protective equipment during patient interaction, these HCWs were further disadvantaged [5]. These challenges were exacerbated by existing socio-demographic disparities such as living in multi-generational households and underlying health conditions [10,11].

In addition to increased COVID-19 contact, we also found an association between ethnicity and the provision of training and supervision during redeployment. Notably, Asian HCWs, particularly doctors, were less likely to report receiving training during redeployment than their White counterparts. This finding is concerning, especially considering the recognised importance of training for successful redeployment during the pandemic [14,15]. A qualitative study conducted among ethnic minority staff in the UK pointed out that in addition to their exclusion from risk assessments, they were also excluded from training discussions, highlighting a lack of support when managers were from non-ethnic minority backgrounds [29]. In contrast, HCWs, especially doctors, from Black ethnic backgrounds were significantly more likely to report receiving supervision compared to their White counterparts. While clinical supervision has various benefits including reduced stress and anxiety, better quality of care delivery, and better working environment [33], it is essential to address any negative connotations associated with supervision among ethnic minority staff, such as being supervised due to a lack of trust in their experience and skills.

The increase in COVID-19 contact among ethnic minority staff, despite being at higher risk, lower reports of changes post-risk assessment, and reduced training during redeployment may be attributed to structural discrimination [31]. Their reluctance to voice concerns about risk assessments [34] and inability to challenge redeployment decisions [25] may potentially stem from past experiences of harassment, bullying or abuse at work [9]. Persistent structural discrimination within the NHS has been well-documented in literature [9,25,35]. The disparities in COVID-19 contact, training and supervision based on ethnicity, as highlighted in our study, warrant further investigation in the context of structural discrimination. Utilizing an intersectional lens will enable a better understanding of how these disparities proliferate.

### Strengths and Limitations

Our sample is diverse, large, and our cohort has been extensively phenotyped, which allows us to look beyond which groups were more likely to be redeployed and to examine experiences of training/supervision and patient contact. Our study also has some limitations. Since it is a cross-sectional analysis, we cannot determine the direction of causality. Further, participants perception of redeployment experiences, might have changed over time, and those with poor experiences were potentially more likely to remember lack of training/ supervision, leading to recall bias. As with any consented cohort study, there may be volunteer bias, however, our cohort closely mirrors the NHS workforce, indicating the representativeness of our sample, albeit with a lower proportion of ancillary staff [36]. Our findings relating to COVID-19 contact during redeployment should be interpreted with caution. While the questionnaire items relating to HCW’s redeployment and their interaction with COVID-19 patients inquire about similar periods (“during the UK national lockdown” for redeployment and “in the first month after the start of UK national lockdown” for COVID-19 interaction), it is possible that participants were redeployed after the first month and thus could have been reporting COVID-19 contact in their usual role. Given that most of the redeployment in the NHS occurred in March and April 2020, we assume that reports of COVID-19 contact relate to the redeployed role [37,38].

## Conclusion

In conclusion, we found associations between ethnicity, migration status and occupational role of healthcare workers with redeployment and post-redeployment experiences. Our study is also the first of its kind to report differences in redeployment experiences based on migration status of HCWs. Future studies should include qualitative work, to capture the complexity of redeployment experiences and the circumstances around participants’ ability to refuse redeployment and analyses of routinely collected NHS human resources data which would not be influenced by reporting and selection bias. It is crucial to address the inequalities highlighted in our study and in previous work by implementing policy to address the impact of racism and structural discrimination in healthcare. This will ultimately foster greater inclusivity of ethnic minority staff and protect them in times of crises within the healthcare system.

## Contributions

The idea for UK-REACH, including the funding application was led by MP with input from KW, and the study collaborative group. The questionnaire design involved CAM, KW, KK, MP, and the study collaborative group. LB developed the online consent and questionnaire tools. This particular analysis idea was formulated by ZL, CAM and MP. ZL conducted the data analysis with input from CAM, MP, and KW, and drafted the manuscript with their input. All authors, MG, IQ, PP, SL, LN, AAO, and JC collectively reviewed, edited, and approved the final manuscript for publication, with MP serving as the guarantor.

## Data availability statement

To access data or samples produced by the UK-REACH study, the working group representative must first submit a request to the□Core Management Group□by contacting the UK-REACH Project Manager in the first instance. For ancillary studies outside of the core deliverables, the□Steering Committee□will make final decisions once they have been approved by the□Core Management Group.□□Decisions on granting the access to data/materials will be made within eight weeks. □

□

Third party requests from outside the Project will require explicit approval of the Steering Committee once approved by the Core Management Group.□

□

Note that should there be significant numbers of requests to access data and/or samples then a separate□Data Access Committee□will be convened to appraise requests in the first instance.

## Data Availability

To access data or samples produced by the UK-REACH study, the working group representative must first submit a request to the Core Management Group by contacting the UK-REACH Project Manager in the first instance. For ancillary studies outside of the core deliverables, the Steering Committee will make final decisions once they have been approved by the Core Management Group.  Decisions on granting the access to data/materials will be made within eight weeks.  

https://uk-reach.org/main/data-dictionary/

## Acknowledgements

We would like to thank all the participants who have taken part in this study when the NHS is under immense pressure. We wish to acknowledge the Professional Expert Panel group (Amir Burney, Association of Pakistani Physicians of Northern Europe; Tiffanie Harrison; London North West University Healthcare NHS Trust; Ahmed Hashim, Sudanese Doctors Association; Sandra Kazembe, University Hospitals Leicester NHS Trust; Susie M. Lagrata (Co-chair), Filipino Nurses Association, UK & University College London Hospitals NHS Foundation Trust; Satheesh Mathew, British Association of Physicians of Indian Origin; Juliette Mutuyimana, Kingston Hospitals NHS Trust; Padmasayee Papineni (Co-chair), London North West University Healthcare NHS Trust; Tatiana Monteiro, University Hospitals Leicester NHS Trust), the UK-REACH Stakeholder Group ^38^, the Study Steering Committee, Serco, as well as the following people and organisations for their support in setting up the study from the regulatory bodies: Kerrin Clapton and Andrew Ledgard (General Medical Council), Caroline Kenny (Nursing and Midwifery Council), David Teeman and Lisa Bainbridge (General Dental Council), My Phan and Jenny Clapham (General Pharmaceutical Council), Angharad Jones (General Optical Council), Mark Neale (Pharmaceutical Society of Northern Ireland) and the Health and Care Professions Council. We would also like to acknowledge the following trusts and sites who recruited participants to the study: Affinity Care, Berkshire Healthcare NHS Trust, Birmingham and Solihull NHS Foundation Trust, Birmingham Community Healthcare NHS Foundation Trust, Black Country Community Healthcare NHS Foundation Trust, Bridgewater Community Healthcare NHS Trust, Central London Community Healthcare NHS Trust, Chesterfield Royal Hospital NHS Foundation Trust, County Durham and Darlington Foundation Trust, Derbyshire Healthcare NHS Foundation Trust, Lancashire Teaching Hospitals NHS Foundation Trust, Lewisham and Greenwich NHS Trust, London Ambulance NHS Trust, NHS Borders, Northumbria Healthcare NHS Foundation Trust, Nottinghamshire Healthcare NHS Foundation Trust, Royal Brompton and Harefield NHS trust, Royal Free NHS Foundation Trust, Sheffield Teaching Hospitals NHS Foundation Trust, South Central Ambulance Service NHS Trust, South Tees NHS Foundation Trust, St George’s University Hospital NHS Foundation Trust, Sussex Community NHS Foundation Trust, University Hospitals Coventry and Warwickshire NHS Trust, University Hospitals of Leicester NHS Trust, University Hospitals Southampton NHS Foundation Trust, Walsall Healthcare NHS Trust and Yeovil District Hospital NHS Foundation Trust.

## References

1. WHO. WHO Coronavirus (COVID-19) Dashboard [online]. 2023. https://covid19.who.int/ [accessed 4 Oct 2023].

2. Aldridge RW, Lewer D, Katikireddi SV, Mathur R, Pathak N, Burns R, et al. Black, Asian and Minority Ethnic groups in England are at increased risk of death from COVID-19: indirect standardisation of NHS mortality data. Wellcome Open Res. 2020 Jun 24;5:88.

3. Raleigh V. The King’s Fund. 2022. Deaths from Covid-19 (coronavirus): how are they counted and what do they show? https://www.kingsfund.org.uk/publications/deaths-covid-19 [accessed 4 Oct 2023].

4. Nguyen LH, Drew DA, Graham MS, Joshi AD, Guo CG, Ma W, et al. Risk of COVID-19 among front-line health-care workers and the general community: a prospective cohort study. Lancet Public Health. 2020 Sep;5(9):e475–83.

5. Martin CA, Pan D, Nazareth J, Aujayeb A, Bryant L, Carr S, et al. Access to personal protective equipment in healthcare workers during the COVID-19 pandemic in the United Kingdom: results from a nationwide cohort study (UK-REACH). BMC Health Serv Res. 2022 Dec 5;22(1):867.

6. Cooper K. BAME doctors hit worse by lack of PPE. British Medical Association [online]. 2020. Available from: https://www.bma.org.uk/news-and-opinion/bame-doctors-hit-worse-by-lack-of-ppe [accessed 4 Oct 2023].

7. Katikireddi SV, Lal S, Carrol ED, Niedzwiedz CL, Khunti K, Dundas R, et al. Unequal impact of the COVID-19 crisis on minority ethnic groups: a framework for understanding and addressing inequalities. J Epidemiol Community Health (1978). 2021 Oct;75(10):970–4.

8. Martin CA, Patel P, Goss C, Jenkins DR, Price A, Barton L, et al. Demographic and occupational determinants of anti-SARS-CoV-2 IgG seropositivity in hospital staff. J Public Health (Bangkok). 2022 Jun 27;44(2):234–45.

9. NHS. NHS Workforce Race Equality Standard. 2022.

10. Pan D, Sze S, Irizar P, George N, Chaka A, Lal Z, et al. Are clinical outcomes from COVID-19 improving in ethnic minority groups? EClinicalMedicine. 2023 Jul;61:102091.

11. Public Health England. Disparities in the risk and outcomes of COVID-19. London; 2020 Aug.

12. Cook Tim, Kursumovic Emira, Lennane Simon. Exclusive: deaths of NHS staff from COVID-19 analysed. 2020. Available from: https://www.hsj.co.uk/exclusive-deaths-of-nhs-staff-from-covid-19-analysed/7027471.article [accessed 30 Nov 2023].

13. Propper C, Stoye G, Zaranko B. The Wider Impacts of the Coronavirus Pandemic on the NHS *. Fisc Stud. 2020 Jun 26;41(2):345–56.

14. Vera San Juan N, Clark SE, Camilleri M, Jeans JP, Monkhouse A, Chisnall G, et al. Training and redeployment of healthcare workers to intensive care units (ICUs) during the COVID-19 pandemic: A systematic review. BMJ Open. 2022 Jan 7;12(1).

15. NHS. COVID-19: Deploying our people safely [online]. 2020. Available from: https://www.england.nhs.uk/coronavirus/documents/covid-19-deploying-our-people-safely/ [accessed 4 Oct 2023].

16. Kennedy E, Kennedy P, Hernandez J, Shakoor K, Munyan K. Understanding Redeployment During the COVID-19 Pandemic: A Qualitative Analysis of Nurse Reported Experiences. SAGE Open Nurs. 2022 Jan 21;8:237796082211149.

17. Sykes A, Pandit M. Experiences, challenges and lessons learnt in medical staff redeployment during response to COVID-19. BMJ Leader. 2021 Jun;5(2):98–101.

18. Kapilashrami A, Otis M, Omodara D, Nandi A, Vats A, Adeniyi O, et al. Ethnic disparities in health & social care workers’ exposure, protection, and clinical management of the COVID-19 pandemic in the UK. Crit Public Health 32. 1–14. 10.1080/09581596.2021.1959020.

19. Gogoi M, Reed-Berendt R, Al-Oraibi A, Hassan O, Wobi F, Gupta A, et al. Ethnicity and COVID-19 outcomes among healthcare workers in the UK: UK-REACH ethico-legal research, qualitative research on healthcare workers’ experiences and stakeholder engagement protocol. BMJ Open. 2021 Jul 9;11(7):e049611.

20. Woolf K, Melbourne C, Bryant L, Guyatt AL, McManus IC, Gupta A, et al. The United Kingdom Research study into Ethnicity And COVID-19 outcomes in Healthcare workers (UK-REACH): protocol for a prospective longitudinal cohort study of healthcare and ancillary workers in UK healthcare settings. BMJ Open. 2021 Sep 17;11(9):e050647.

21. Office for National Statistics. Ethnic group, England and Wales: Census 2021 [online]. 2022. Available from: https://www.ons.gov.uk/peoplepopulationandcommunity/culturalidentity/ethnicity/bulletins/ethnicgroupenglandandwales/census2021 [accessed17 Jan 2024]

22. Ministry of Housing C& LG. English indices of deprivation 2019. 2019 Sep.

23. UK Health Security Agency and Department of Health and Social Care. COVID-19: guidance for people whose immune system means they are at higher risk. 2023 Sep.

24. Rubin DB. Inference and missing data. Biometrika. 1976;63(3):581–92.

25. Rhead R, Harber-Aschan L, Onwumere J, Polling C, Dorrington S, Ehsan A, et al. Ethnic inequalities among NHS staff in England - workplace experiences during the COVID-19 pandemic. Available from: 10.1101/2023.04.13.23288481

26. Martin CA, Pan D, Melbourne C, Teece L, Aujayeb A, Baggaley RF, et al. Risk factors associated with SARS-CoV-2 infection in a multiethnic cohort of United Kingdom healthcare workers (UK-REACH): A cross-sectional analysis. PLoS Med. 2022 May 1;19(5).

27. Cai H, Tu B, Ma J, Chen L, Fu L, Jiang Y, et al. Psychological impact and coping strategies of frontline medical staff in Hunan between January and March 2020 during the outbreak of coronavirus disease 2019 (COVID) in Hubei, China. Medical Science Monitor. 2020 Apr 15;26.

28. Ballantyne H, Achour N. The Challenges of Nurse Redeployment and Opportunities for Leadership During COVID-19 Pandemic. Disaster Med Public Health Prep. 2023 Feb 14;17(15–16).

29. Jesuthasan J, Powell RA, Burmester V, Nicholls D. “We weren’t checked in on, nobody spoke to us”: An exploratory qualitative analysis of two focus groups on the concerns of ethnic minority NHS staff during COVID-19. BMJ Open. 2021 Dec 31;11(12).

30. Qureshi I, Pareek M, Gogoi M, Wobi F, Chaloner J, Al-Oraibi A, et al. Healthcare Workers From Diverse Ethnicities and Their Perceptions of Risk and Experiences of Risk Management During the COVID-19 Pandemic: Qualitative Insights From the United Kingdom-REACH Study. 2022; Frontiers in medicine, 9, 930904. 10.3389/fmed.2022.930904

31. Martin CA, Woolf K, Bryant L, Goss C, Gogoi M, Lagrata S, et al. Coverage, completion and outcomes of COVID-19 risk assessments in a multi-ethnic nationwide cohort of UK healthcare workers: a cross-sectional analysis from the UK-REACH Study. Occup Environ Med. 2023 Jul;80(7):399–406.

32. Iacobucci G. Covid-19: Many trusts have not done risk assessments for ethnic minority staff, *BMJ* investigation finds. BMJ. 2020 Jul 10;m2792.

33. Health & care professions council. The benefits and outcomes of effective supervision [Internet]. 2021. Available from: https://www.hcpc-uk.org/standards/meeting-our-standards/supervision-leadership-and-culture/supervision/the-benefits-and-outcomes-of-effective-supervision/ [accessed 27 Nov 2023].

34. Iacobucci G. Covid-19: Many trusts have not done risk assessments for ethnic minority staff, *BMJ* investigation finds. BMJ. 2020 Jul 10;m2792.

35. Silverio SA, De Backer K, Dasgupta T, Torres O, Easter A, Khazaezadeh N, et al. On race and ethnicity during a global pandemic: An ‘imperfect mosaic’ of maternal and child health services in ethnically-diverse South London, United Kingdom. EClinicalMedicine. 2022 Jun;48:101433.

36. Woolf K, McManus IC, Martin CA, Nellums LB, Guyatt AL, Melbourne C, et al. Ethnic differences in SARS-CoV-2 vaccine hesitancy in United Kingdom healthcare workers: Results from the UK-REACH prospective nationwide cohort study. The Lancet Regional Health - Europe. 2021 Oct;9:100180.

37. Royal College of Physicians. Tracking the impact of COVID-19 on the workforce [online]. 2020. Available from: https://www.rcplondon.ac.uk/news/tracking-impact-covid-19-workforce [accessed 27 Nov 2023]

38. Willan J, King AJ, Jeffery K, Bienz N. Challenges for NHS hospitals during covid-19 epidemic. BMJ. 2020 Mar 20;m1117.

