## Supplementary data for "Redeployment Experiences of Healthcare Workers in the UK during COVID-19: data from the nationwide UK-REACH study"

On behalf of the UK-REACH Study Collaborative Group:

Manish Pareek (Chief investigator, University of Leicester), Laura Gray (University of Leicester), Laura Nellums (University of New Mexico), Anna L Guyatt (University of Leicester), Catherine John (University of Leicester), I Chris McManus (University College London), Katherine Woolf (University College London), Ibrahim Abubakar (University College London), Amit Gupta (Oxford University Hospitals), Avinash Aujayeb (Northumbria Specialist Emergency Care Hospital), Bindu Gregary (Royal Preston Hospital), Rubina Reza (Derbyshire Healthcare NHS Foundation Trust), Sandra Simpson (Nottinghamshire Healthcare NHS Foundation Trust), Stephen Zingwe (Berkshire Healthcare NHS Foundation Trust), Keith R Abrams (University of York), Martin D Tobin (University of Leicester), Louise Wain (University of Leicester), Sue Carr (University Hospitals of Leicester NHS Trust),  Edward Dove (University of Edinburgh),  Kamlesh Khunti (University of Leicester), David Ford (University of Swansea), Robert Free (University of Leicester).

**Supplementary Text 1. UK-REACH – overview and recruitment**

**Overview**

UK-REACH, a nationwide research initiative comprising multiple sub-studies, is dedicated to understanding the degree of impact of the COVID-19 pandemic on healthcare workers (HCWs) from ethnic minority backgrounds.

This work utilizes data from the baseline questionnaire of the cohort study, administered between December 2020 and March 2021. The study design, sampling and measures included in the questionnaire are found in the study protocol^19^, the cohort profile^18^, the data dictionary, <https://www.uk-reach.org/data-dictionary>, and in other published work^5,24,30,33^.

**Recruitment**

Recruitment efforts targeted participants aged 16 and above, residing in the UK, and working as HCWs or ancillary workers in a healthcare setting and/ or registered with one of the seven major UK professional regulatory bodies:

1. The General Medical Council (GMC)
2. The Nursing and Midwifery Council (NMC)
3. The General Dental Council (GDC)
4. The Health and Care Professions Council (HCPC)
5. The General Optical Council (GOC)
6. The General Pharmaceutical Council (GPC)
7. The Pharmaceutical Society of Northern Ireland (PSNI)

Professional regulatory bodies facilitated recruitment by sending emails to their registrants containing a hyperlink to the study website. Interested individuals could create a user profile, review the participant information sheet, and, upon willingness, electronically sign a consent form, Subsequently, participants were prompted to complete the online questionnaire. The sample size was augmented by recruiting HCWs through participating healthcare trusts, along with promotional efforts on social media and in newsletters. These additional participants underwent the same recruitment process.

Participation rates at each stage adhere to the guidelines outlined in the Checklist for Reporting Results of Internet E-Surveys (CHERRIES).

**Supplementary Text 2. Involvement and engagement**

A Professional Expert Panel of HCWs from a range of ethnic backgrounds, healthcare occupations, sexes, and national and local organisations worked with us closely on the project. They were involved in the design of the survey instruments and assisted in developing the research question and analysis plan. Some members have critically reviewed the manuscript and are co-authors of the study.

**Supplementary Table 1. Derivation of variables used in the analysis**

| **Variable** | **Description** |
| --- | --- |
| **Ethnicity** | Categorical variable. Participants were asked to select their ethnicity from a list of the 18 Office for National Statistics categories:  Asian/Asian British – Indian Asian/Asian British – Pakistani  Asian/Asian British – Bangladeshi Asian/Asian British – Chinese Asian/Asian British - Any other Asian background Black/African/Caribbean/Black British - African Black/African/Caribbean/Black British – Caribbean Black/African/Caribbean/Black British - Any other Black/African/Caribbean background Mixed/Multiple ethnic groups - White and Black Caribbean Mixed/Multiple ethnic groups - White and Black African Mixed/Multiple ethnic groups - White and Asian Mixed/Multiple ethnic groups - Any other Mixed/multiple ethnic background White - English/Welsh/Scottish/Northern Irish/British White – Irish White - Gypsy or Irish Traveller White - Any other white background Other ethnic group – Arab Other ethnic group - Any other ethnic background  These were categorised into the 5 broader Office for National Statistics ethnicity categories (Asian, Black, Mixed, White, Other). |
| **Age** | Categorical variable. Age in years. Derived from date of birth entered by participants at registration |
| **Sex** | Binary variable. Participants were asked their sex assigned at birth. |
| **Migration Status** | Binary variable. Born in the UK vs Born overseas. Participants were asked whether they were born in the UK. |
| **Occupation** | Categorical variable. Participants were asked to select their main job role. **Categorised as below:**  **Doctors –** Doctor    **Nurse, NA or Midwife –** Advanced Nurse Practitioner, Healthcare assistant, Maternity support worker, Midwife, Nurse, Nursing associate, other nursing and midwifery role.    **Allied Health Professional –** arts therapist, chiropodist/ podiatrist, dietician, Hearing aid dispenser, Occupational therapist, Operating department practitioner, Orthoptist, Physiotherapist, Practitioner psychologist, Prosthetist/ Orthotist, Radiographer, OT support Worker, Phlebotomist, Physiotherapy assistant, Radiography assistant, other clinical support role.    **Pharmacy –** pharmacy, pharmacy technician, and other pharmacy role.    **Healthcare scientist –** biomedical scientist, clinical scientist    **Ambulance –** emergency medical technician, paramedic, other ambulance role    **Dental –** clinical dental technician, dental hygienist, dental nurse, dental technician, dentist, other dental role    **Optical –** dispensing optician, optometrist, other optical role    **Administrative –** administration    **Estates/ facilities –** catering services, domestic services, estates services, porter, and other wider healthcare roles.    **Other –** Medical associates including Advanced Critical Care Practitioner, Anaesthesia associate, Surgical Care Practitioner, Physician associate, other medical associates, and any other role. |
| **Index of Multiple Deprivation** | Ordinal variable. Participants provided their residential postcode on registration for the study. This was used to determine the Index of Multiple Deprivation (the official measure of deprivation for small areas of England) in the area in which they live. The IMD ranks all areas in England based on 7 measures of deprivation and the ranks can be expressed as quintiles. Lower quintiles indicate more deprivation. Although Wales, Scotland and Northern Ireland have their own measures of deprivation, these are said not to be directly comparable to English IMD and therefore we elected to impute an ‘English IMD’ for residents of the these nations. |
| **Underlying health conditions** | Categorical variable (ordered). Number of long-term physical or mental health conditions from a selection of diabetes, heart disease, hypertension, asthma, other lung conditions, kidney or liver disease, neurological disease, cancer, immunosuppression, organ transplant, depression, anxiety, other psychiatric disorders. |
| **Job Sector** | Categorical variable. Participants were asked which sector they worked in during the lockdown. This was categorised into: “1, NHS \| 2, Other public sector (e.g. local or national government) \| 3, Private sector \| 4, Private facility temporarily used by the NHS \| 5, University/ higher education \| 99, Prefer not to answer” |

For further information on questionnaire variables, refer to the UK-REACH data dictionary (<https://www.uk-reach.org/data-dictionary>).

**Supplementary Table 2: Seniority level of HCWs within the analysed cohort:**

| Job bands: |  |
| --- | --- |
| Band 1-4 | 472 (4.3) |
| Band 5 | 1171 (10.8) |
| Band 6 | 2029 (18.6) |
| Band 7 | 1476 (13.6) |
| Band 8 & 9 | 901 (8.3) |
| Missing | 4840 (44.5) |

| Doctor grade: |  |
| --- | --- |
| Foundation Doctor | 144 (1.3) |
| Core & Speciality Trainee | 538 (5.0) |
| Locally employed/ trust doctor | 211 (1.94) |
| GP | 555 (5.1) |
| Consultant | 966 (8.9) |
| SAS | 182 (1.7) |
| Other | 63 (0.6) |
| Missing | 8230 (75.6) |

**Supplementary Table 2** describes the cohort based on their level of seniority. Agenda for change (AfC) pay band and doctors’ grades are proxy measures for occupational seniority. Band 5, the reference category, corresponds to the salary level of a newly qualified nurse. In the analysis of doctors, a foundational trainee refers to a newly qualified doctor within first two years of training after medical school. A core trainee has completed foundational training and chosen a broad area of specialisation. A speciality trainee, also known as a registrar, is undergoing training in a specific speciality. A consultant has completed training in a specific speciality, and a GP has completed training in general practice and typically practices in the community rather than in a hospital setting. All data in the right-hand column are n (%).

**Supplementary Table 3. Univariable analysis of factors associated with redeployment during COVID-19**

| **Variable** | **Not redeployed 8667 (79.6)** | **Redeployed**  **2223 (20.4)** | **Unadjusted OR**  **(95% CI)** | **P value** |
| --- | --- | --- | --- | --- |
| **Demographic and health factors** | | | | |
| **AGE** |  | | | |
| <30 years | 1176 (73.7) | 420 (26.3) | Ref |  |
| 31-45 years | 3146 (78.9) | 841 (21) | 0.75 (0.65 – 0.86) | <0.001 |
| 46-60 years | 3500 (81) | 819 (19) | 0.66 (0.57 – 0.75) | <0.001 |
| 61 and above | 800 (85.8) | 133 (14.3) | 0.47 (0.37 – 0.58) | <0.001 |
| **SEX** |  | | | |
| Male | 2110 (78.9) | 566 (21.2) | Ref  0.94 (0.85 – 1.05) | 0.29 |
| Female | 6533 (79.8) | 1654 (20.2) |  |  |
| **ETHNICITY** |  | | | |
| White | 6127 (79.8) | 1556 (20.3) | Ref | Ref |
| Black | 376 (81.7) | 84 (18.3) | 0.88 (0.69 – 1.12) | 0.3 |
| Asian | 1652 (79.6) | 424 (20.4) | 1.01 (0.9 – 1.14) | 0.86 |
| Mixed | 338 (75.5) | 110 (24.6) | 1.28 (1.03 – 1.6) | 0.03 |
| Other | 173 (77.9) | 49 (22) | 1.12 (0.81 – 1.54) | 0.5 |
| **MIGRATION STATUS** |  | | | |
| Born in the UK | 6394 (79.9) | 1606 (20) | Ref |  |
| Born abroad | 2272 (78.6) | 617 (21.4) | 1.08 (0.97 – 1.2) | 0.14 |
| **INDEX OF MULTIPLE DEPRIVATION (quintile)** |  | | | |
| 1 (most deprived) | 733 (77.1) | 212 (22.9) | 1.17 (0.97 – 1.42) | 0.09 |
| 2 | 1282 (78.9) | 342 (21.1) | 1.05 (0.9 – 1.24) | 0.5 |
| 3 | 1573 (79.8) | 398 (20.2) | Ref | Ref |
| 4 | 1858 (79.4) | 481 (20.6) | 1.02 (0.88 – 1.19) | 0.76 |
| 5 (least deprived) | 2185 (79.7) | 555 (20.3) | 1.00 (0.87 – 1.16) | 0.96 |
| **COMORBIDITIES** |  | | | |
| No conditions | 3621 (79.2) | 952 (20.8) | Ref  0.95 (0.84 – 1.08) | Ref |
| Long-term conditions that required shielding during COVID-19 | 1685 (80.0) | 422 (20.0) |  | 0.46 |
| Other conditions | 2597 (80.4) | 635 (19.7) | 0.93 (0.83 – 1.04) | 0.21 |
| **Occupational factors** | | | | |
| **OCCUPATION** |  |  |  |  |
| Doctors | 2101 (78.2) | 587 (22) | Ref | Ref |
| Nurses, NAs, Midwives | 1921 (77.3) | 564 (22.7) | 1.05 (0.92 – 1.2) | 0.46 |
| Allied Health Professionals | 2551 (77.1) | 756 (22.9) | 1.06 (0.94 – 1.2) | 0.34 |
| Pharmacy | 209 (89.7) | 24 (10.3) | 0.41 (0.27 - 0.63) | <0.001 |
| Healthcare scientist | 475 (89.3) | 57 (10.7) | 0.43 (0.32 – 0.57) | <0.001 |
| Ambulance | 388 (87.8) | 54 (12.2) | 0.5 (0.37 – 0.67) | <0.001 |
| Dental | 360 (82.8) | 75 (17.2) | 0.75 (0.57 – 0.97) | 0.03 |
| Optical | 102 (87.2) | 15 (13) | 0.53 (0.30 – 0.91) | 0.02 |
| administrative | 201 (88.6) | 26 (11.5) | 0.46 (0.30 – 0.70) | <0.001 |
| Estates/ facilities | 78 (75.7) | 25 (24.3) | 1.15 (0.72 – 1.82) | 0.56 |
| Other (including medical associates) | 280 (87.5) | 40 (12.5) | 0.51 (0.36 – 0.72) | <0.001 |
| **JOB SECTOR** | | | | |
| Worked for the NHS in some capacity | 7037 (77.7) | 2019 (22.3) | 2.71 (2.25 – 3.26) | <0.001 |
| Worked outside the NHS | 1254 (90.4) | 133 (9.6) | Ref | Ref |

**Supplementary Table 3** shows the results of univariable logistic regression as odds ratios, 95% confidence intervals and p values for the association of socio-demographic, health and occupational variables with outcome 1 (reporting redeployment).

In the occupation variable, medical associates were moved to the ‘other’ category, to retain sufficient statistical power

**Supplementary Table 4: Univariable analysis of job bands and doctor grades with redeployment:**

| SENIORITY LEVEL: | Not redeployed | Redeployed | Unadjusted OR (95% CI) | P value |
| --- | --- | --- | --- | --- |
| JOB BAND |  |  |  |  |
| Band 1-4 | 381 (81) | 91 (19.3) | Ref | Ref |
| Band 5 | 859 (73.4) | 312 (26.6) | 1.52 (1.17 – 1.98) | 0.002 |
| Band 6 | 1547 (76.2) | 483 (23.8) | 1.31 (1.02 – 1.68) | 0.04 |
| Band 7 | 1137 (77) | 338 (23) | 1.25 (0.96 – 1.62) | 0.09 |
| Band 8 & 9 | 731 (81.1) | 170 (19) | 0.97 (0.73 – 1.3) | 0.85 |
| DOCTOR GRADE |  |  |  |  |
| Foundation doctor | 64 (44.4) | 80 (55.6) | Ref | Ref |
| Core & speciality trainee |  |  | 0.28 (0.2 – 0.41) | <0.001 |
| Locally employed/ trust doctor | 152 (72) | 59 (28) | 0.31 (0.2 – 0.48) | <0.001 |
| GP | 506 (91.2) | 49 (8.8) | 0.08 (0.05 – 0.12) | <0.001 |
| Consultant | 758 (78.5) | 208 (21.5) | 0.22 (0.15 – 0.32) | <0.001 |
| SAS | 149 (81.9) | 33 (18.1) | 0.18 (0.11 – 0.29) | <0.001 |
| Other | 49 (77.8) | 14 (22.2) | 0.23 (0.12 – 0.45) | <0.001 |

**Supplementary Table 4** shows the results of univariable logistic regression as odds ratios, 95% confidence intervals and p values for the association of job bands and doctor grades with outcome 1 (redeployment). In this analysis, we adjust for the grade or stage of training for doctors and for the agenda for change pay band for HCWs on these pay scales. The NHS agenda for change pay bands comprise bands 1-9 (with salary increasing as band level rises) for HCWs other than doctors, dentists and very senior managers. This scale was used as a proxy measure for occupational seniority.

**Supplementary Table 5: Multivariable analysis of ethnicity, migration status, and occupation with redeployment, training, supervision, patient contact and COVID-19 contact after adjustment for demographic and health covariates**

| Variable | Redeployment |  | Training |  | Supervision |  | Patient Contact |  | COVID-19 contact |  |
| --- | --- | --- | --- | --- | --- | --- | --- | --- | --- | --- |
| Demographic and household factors | **aOR** | **P value** | aOR | P value | aOR | P value | aOR | P value |  | P value |
| ETHNICITY |  |  |  |  |  |  |  |  |  |  |
| White | Ref | Ref | Ref | Ref | ref | ref | ref | Ref | ref | Ref |
| Black/ African/ Caribbean/ Black | 0.8 (0.62 – 1.04) | 0.1 | 0.99 (0.58 – 1.68) | 0.97 | 2.02 (1.14 – 3.57) | 0.02 | 0.71 (0.43 – 1.18) | 0.18 | 1.33 (1.07 - 1.66) | 0.009 |
| Asian/ Asian British | 0.89 (0.76 – 1.02) | 0.1 | 0.66 (0.5 – 0.88) | 0.005 | 0.89 (0.66 – 1.2) | 0.42 | 1.16 (0.84– 1.59) | 0.36 | 1.30 (1.14 - 1.48) | <0.001 |
| Mixed/ multiple | 1.21 (0.96 – 1.52) | 0.10 | 1 (0.65 – 1.53) | 0.99 | 1.35 (0.86 – 2.12) | 0.2 | 0.87 (0.56 – 1.36) | 0.55 | 0.93 (0.76 - 1.16) | 0.53 |
| Other | 0.99 (0.7 – 1.39) | 0.94 | 0.86 (0.46 – 1.6) | 0.63 | 1.01 (0.53 – 1.92) | 0.97 | 1.58 (0.75 – 3.35) | 0.23 | 1.28 (0.94 - 1.74) | 0.11 |
| MIGRATION |  |  |  |  |  |  |  |  |  |  |
| Born in the UK | Ref | ref | Ref | ref | ref | Ref | Ref | Ref | Ref | Ref |
| Born outside the UK | 1.11 (0.98 -1.26) | 0.1 | 1.30 (1.01 - 1.66) | 0.04 | 1.06 (0.83 – 1.35) | 0.64 | 0.99 (0.76 – 1.28) | 0.93 | 1.27 (1.13 - 1.42) | <0.001 |
| AGE |  |  |  |  |  |  |  |  |  |  |
| 0-31 years | Ref | Ref | ref | Ref | Ref | ref | ref | ref | ref | ref |
| 31-45 years | 0.73 (0.63 – 0.84) | <0.001 | 1.11 (0.86 -1.44) | 0.4 | 0.7 (0.54 – 0.92) | 0.01 | 0.62 (0.46 – 0.83) | 0.002 | 0.66 (0.58 – 0.75) | <0.001 |
| 46-60 years | 0.63 (0.55 – 0.73) | <0.001 | 1.02 (0.79 – 1.32) | 0.87 | 0.53 (0.4 – 0.7) | <0.001 | 0.39 (0.29 – 0.53) | <0.001 | 0.41 (0.36 – 0.47) | <0.001 |
| 60 - max | 0.47 (0.38 – 0.59) | <0.001 | 1.3 (0.84 – 2.01) | 0.24 | 0.53 (0.34 – 0.82) | 0.004 | 0.28 (0.18 – 0.44) | <0.001 | 0.24 (0.20 – 0.29) | <0.001 |
| SEX |  |  |  |  |  |  |  |  |  |  |
| Male | Ref | ref | Ref | ref | ref | Ref | ref | Ref | ref | Ref |
| Female | 0.82 (0.73 – 0.93) | 0.001 | 0.92 (0.74 – 1.15) | 0.48 | 0.91 (0.72 – 1.14) | 0.41 | 1.15 (0.91 – 1.46) | 0.25 | 0.70 (0.63 – 0.77) | <0.001 |
| DEPRIVATION |  |  |  |  |  |  |  |  |  |  |
| 1 (most deprived) | 1.09 (0.9 – 1.32) | 0.36 | 0.99 (0.69 – 1.42) | 0.97 | 0.96 (0.67 – 1.39) | 0.85 | 0.87 (0.58 – 1.28) | 0.47 | 1.19 (1.00 – 1.42) | 0.05 |
| 2 | 1.04 (0.88 – 1.22) | 0.64 | 0.8 (0.58 – 1.1) | 0.16 | 0.8 (0.58 – 1.09) | 0.15 | 0.91 (0.66 – 1.27) | 0.58 | 1.15 (0.99 – 1.34) | 0.08 |
| 3 | Ref | Ref | Ref | ref | Ref | ref | ref | Ref | ref | Ref |
| 4 | 1.05 (0.9 – 1.22) | 0.53 | 0.94 (0.71 – 1.25) | 0.68 | 0.91 (0.70 – 1.23) | 0.61 | 1.06 (0.77 – 1.45) | 0.72 | 0.98 (0.85 - 1.13) | 0.77 |
| 5 (least deprived) | 1.04 (0.9 – 1.21) | 0.56 | 0.84 (0.63 – 1.11) | 0.22 | 0.71 (0.54 – 0.95) | 0.02 | 0.87 (0.65 – 1.16) | 0.34 | 0.91 (0.80 – 1.04) | 0.18 |
| COMORBIDITIES |  |  |  |  |  |  |  |  |  |  |
| No conditions | ref | Ref | *(not relevant for training and supervision)* |  | *(not relevant for training and supervision)* |  | ref | Ref | ref | Ref |
| Long-term conditions that required shielding during COVID-19 | 0.99 (0.84 – 1.07) | 0.39 |  |  |  |  | 0.62 (0.48 – 0.79) | <0.001 | 0.83 (0.74 – 0.94) | <0.003 |
| Other conditions | 0.95 (0.86 – 1.13) | 0.87 |  |  |  |  | 1.05 (0.83 – 1.34) | 0.68 | 0.94 (0.85 – 1.05) | 0.28 |
| Occupational factors |  | | |  |  |  |  |  |  |  |
| OCCUPATION |  |  |  |  |  |  |  |  |  |  |
| Doctors | Ref | Ref | Ref | ref | ref | ref | ref | ref | ref | ref |
| Nursing | 1.22 (1.04 – 1.42) | 0.009 | 1.21 (0.92 – 1.59) | 0.17 | 0.63 (0.48 – 0.83) | 0.001 | 1.52 (1.1 – 2.08) | 0.01 | 1.10 (0.97 – 1.25) | 0.15 |
| Allied Health Professionals (not including scientists) | 1.23 (1.07 – 1.41) | 0.003 | 1.61 (1.25 – 2.1) | <0.001 | 1.11 (0.85 – 1.45) | 0.43 | 0.75 (0.57 – 0.99) | 0.04 | 0.48 (0.42 – 0.54) | <0.001 |
| Pharmacy | 0.52 (0.34 – 0.81) | 0.004 | 1.51 (0.63 – 3.66) | 0.36 | 0.34 (0.14 – 0.81) | 0.02 | 0.71 (0.29 – 1.74) | 0.46 | 0.43 (0.33 – 0.58) | <0.001 |
| Healthcare scientist | 0.44 (0.33 – 0.59) | <0.001 | 3.32 (1.63 – 6.79) | 0.001 | 2.52 (1.21 – 5.35) | 0.02 | 0.64 (0.35 – 1.17) | 0.15 | 0.04 (0.35 – 1.17) | <0.001 |
| Ambulance | 0.46 (0.33 – 0.62) | <0.001 | 1.1 (0.61 – 2) | 0.75 | 0.42 (0.23 – 0.75) | 0.004 | 0.53 (0.29 – 0.98) | 0.04 | 2.14 (0.03 – 0.07) | <0.001 |
| Dental | 0.85 (0.64 – 1.1) | 0.23 | 1.98 (1.15 – 3.41) | 0.01 | 0.72 (0.43 – 1.2) | 0.2 | 0.19 (0.11 – 0.32) | <0.001 | 0.10 (0.07 – 0.13) | <0.001 |
| Optical | 0.81 (0.47 – 1.44) | 0.47 | 0.82 (0.28 – 2.35) | 0.71 | 0.2 (0.06 – 0.65) | 0.007 | 0.13 (0.04 – 0.44) | 0.001 | 0.12 (0.07 – 0.2) | <0.001 |
| Administrative | 0.46 (0.3 – 0.7) | <0.001 | 1.31 (0.56 – 3.06) | 0.53 | 1.81 (0.65 – 5.08) | 0.26 | 0.74 (0.3 – 1.81) | 0.51 | 0.07 (0.05 – 0.12) | <0.001 |
| Estates/ facilities | 1.29 (0.8 – 2.03) | 0.29 | 3.35 (1.11 – 10.07) | 0.03 | 0.97 (0.42 – 2.29) | 0.95 | 1.51 (0.54 – 4.22) | 0.43 | 0.23 (0.14 – 0.38) | <0.001 |
| Other (including medical associates) | 0.61 (0.43 – 0.87) | 0.006 | 1.43 (0.71 – 2.87) | 0.32 | 0.48 (0.24 – 0.96) | 0.04 | 1.16 (0.55 – 2.44) | 0.71 | 0.26 (0.19 – 0.34) | <0.001 |
| JOB SECTOR |  |  |  |  |  |  |  |  |  |  |
| Working within the NHS | 2.68 (2.2 – 3.22) | <0.001 | 0.86 (0.58 – 1.28) | 0.47 | 0.87 (0.58 – 1.3) | 0.5 |  |  |  |  |
| Working outside the NHS | Ref | ref | ref | ref | ref | ref |  |  |  |  |

**Supplementary Table 5** shows the results of multivariable logistic regression as adjusted odds ratios, 95% confidence intervals and p values for the association of socio-demographic and occupational variables with outcome 1 (whether or not a HCW was redeployed to a different role because of the pandemic), outcome 2 (whether or not a HCW received training after redeployment), outcome 3 (whether or not a HCW received supervision after redeployment), outcome 4 (whether direct patient contact stayed the same or decreased vs increased in HCW’s redeployed role), and outcome 5 (interaction with number of confirmed or suspected COVID-19 cases).

In the occupation variable, nursing includes midwives and nursing associates; HCW, healthcare workers; NHS, National Health Service.

**Supplementary Table 6. Subcohort analysis of the relationship between seniority and job role (for doctors) with redeployment, training, supervision, patient contact, and COVID-19 contact:**

| Variable | Redeployment |  | Training |  | Supervision |  | Patient Contact |  | COVID-19 Contact |  |
| --- | --- | --- | --- | --- | --- | --- | --- | --- | --- | --- |
| DOCTOR GRADE | **aOR** | **P value** | **aOR** | **P value** | **aOR** | **P value** | **aOR** | **P**  **value** | **aOR** | **P value** |
| ETHNICITY |  |  |  |  |  |  |  |  |  |  |
| White | ref | ref | ref | ref | ref | ref | ref | ref | ref | ref |
| Black | 0.58 (0.37 – 0.92) | 0.02 | 0.87 (0.34 – 2.20) | 0.77 | 5.41 (1.10 – 26.60) | 0.04 | 0.51 (0.20 – 1.29) | 0.15 | 0.98 (0.68 – 1.43) | 0.93 |
| Asian | 0.89 (0.70 – 1.14) | 0.37 | 0.55 (0.34 – 0.87) | 0.49 | 0.66 (0.39 – 1.14) | 0.67 | 1.20 (0.70 – 2.07) | 0.73 | 1.14 (0.92 – 1.41) | 0.23 |
| Mixed | 0.95 (0.64 – 1.42) | 0.82 | 0.78 (0.39 – 1.57) | 0.01 | 1.21 (0.50 – 2.96) | 0.14 | 0.87 (0.39 – 1.95) | 0.51 | 0.92 (0.65 – 1.31) | 0.65 |
| Other | 0.75 (0.46 – 1.23) | 0.26 | 0.42 (0.17 – 1.01) | 0.05 | 0.57 (0.20 – 1.65) | 0.30 | 1.36 (0.44 – 4.18) | 0.60 | 1.12 (0.73 – 1.73) | 0.60 |
| MIGRATION STATUS |  |  |  |  |  |  |  |  |  |  |
| Born in the UK | Ref | Ref | Ref | Ref | Ref | Ref | Ref | Ref | Ref | Ref |
| Born abroad | 0.92 (0.73 – 1.16) | 0.47 | 1.64 (1.05 – 2.56) | 0.03 | 1.23 (0.73 – 2.06) | 0.44 | 1.37 (0.82 – 2.31) | 0.23 | 1.40 (1.14 – 1.72) | 0.001 |
| AGE |  |  |  |  |  |  |  |  |  |  |
| 0-31 years | Ref | ref | Ref | Ref | Ref | Ref | Ref | Ref | Ref | Ref |
| 31-45 years | 0.87 (0.62 – 1.23) | 0.44 | 2.05 (1.10 – 3.83) | 0.02 | 0.48 (0.18 – 1.31) | 0.15 | 1.25 (0.58 – 2.74) | 0.57 | 0.83 (0.59 – 1.17) | 0.29 |
| 46-60 years | 0.74 (0.49 – 1.22) | 0.16 | 1.63 (0.74 – 3.59) | 0.22 | 0.44 (0.14 – 1.38) | 0.16 | 0.41 (0.16 – 1.06) | 0.07 | 0.51 (0.34 – 0.75) | 0.001 |
| 60 - max | 0.55 (0.33 – 0.93) | 0.03 | 1.76 (0.65 – 4.73) | 0.26 | 0.22 (0.06 – 0.79) | 0.02 | 0.29 (0.09 – 0.94) | 0.04 | 0.21 (0.13 – 0.33) | <0.001 |
| SEX |  |  |  |  |  |  |  |  |  |  |
| Male | Ref | Ref | Ref | Ref | Ref | Ref | Ref | Ref | Ref | Ref |
| Female | 0.83 (0.68 – 1.02) | 0.07 | 0.95 (0.66 – 1.37) | 0.78 | 0.94 (0.61 – 1.44) | 0.76 | 1.22 (0.80 – 1.87) | 0.36 | 0.64 (0.53 – 1.72) | 0.001 |
| DEPRIVATION |  |  |  |  |  |  |  |  |  |  |
| 1 (most deprived) | 0.82 (0.54 – 1.24) | 0.36 | 0.69 (0.33 – 1.46) | 0.34 | 0.50 (0.19 – 1.29) | 0.15 | 0.99 (0.41 - 2.40) | 0.98 | 1.11 (0.74 – 1.64) | 0.62 |
| 2 | 0.80 (0.57 – 1.12) | 0.20 | 0.88 (0.47 – 1.64) | 0.69 | 0.53 (0.24 – 1.16) | 0.11 | 1.71 (0.78 - 3.77) | 0.18 | 1.11 (0.81 – 1.52) | 0.52 |
| 3 | Ref | Ref | Ref | Ref | Ref | Ref | Ref | Ref | Ref | Ref |
| 4 | 0.85 (0.63 – 1.16) | 0.32 | 0.94 (0.55 – 1.60) | 0.83 | 0.73 (0.39 – 1.36) | 0.32 | 1.62 (0.80 – 3.27) | 0.18 | 1.21 (0.93 – 1.58) | 0.15 |
| 5 (least deprived) | 0.88 (0.66 – 1.17) | 0.37 | 0.55 (0.33 – 0.91) | 0.02 | 0.58 (0.32 – 1.06) | 0.08 | 1.80 (0.96 – 3.37) | 0.07 | 1.13 (0.89 – 1.47) | 0.32 |
| COMORBIDITIES |  |  |  |  |  |  |  |  |  |  |
| No conditions | Ref | Ref | *(not relevant for training and supervision)* | | | | Ref | Ref | Ref | Ref |
| Long-term conditions that required shielding during COVID-19 | 1.17 (0.89 – 1.53) | 0.27 |  |  |  |  | 0.61 (0.36 – 1.03) | 0.07 | 0.92 (0.73 – 1.15) | 0.29 |
| Other conditions | 1.03 (0.80 – 1.32) | 0.83 |  |  |  |  | 1.25 (0.73 – 2.15) | 0.41 | 0.89 (0.73 – 1.10) | 0.45 |
| DOCTOR GRADE |  |  |  |  |  |  |  |  |  |  |
| Foundation doctor | 3.54 (2.15 – 5.83) | <0.001 | 1.42 (0.6 – 3.34) | 0.43 | 2.79 (0.77 – 8.03) | 0.09 | 1.39 (0.46 – 4.23) | 0.56 | 1.90 (1.09 - 3.30) | 0.02 |
| Core and specialty trainee | 1.12 (0.82 – 1.53) | 0.47 | 1.47 (0.82 – 2.63) | 0.2 | 12.95 (5.6 – 27.77) | <0.001 | 0.82 (0.39 – 1.7) | 0.59 | 1.15 (0.87 - 1.53) | 0.32 |
| Locally employed/ trust doctor | 1.35 (0.9 – 1.98 | 0.13 | 1.08 (0.51 – 2.26) | 0.84 | 2.93 (1.19 – 6.38) | 0.01 | 1.16 (0.44 – 3.07) | 0.77 | 1.57 (1.07 - 2.31) | 0.02 |
| GP | 0.34 (0.25 – 0.48) | <0.001 | 2.78 (1.34 – 5.78) | 0.006 | 1.34 (0.67 – 2.53) | 0.39 | 0.27 (0.13 – 0.56) | <0.001 | 0.46 (0.36 - 0.57) | <0.001 |
| Consultant | Ref | ref | Ref | ref | Ref | Ref | Ref | ref | Ref | Ref |
| SAS | 0.86 (0.57 – 1.3) | 0.47 | 1.03 (0.47 – 2.27) | 0.93 | 3.14 (1.33 – 7.42) | 0.009 | 1. 89 (0.69 – 5.09) | 0.22 | 0.87 (0.62 - 1.22) | 0.42 |
| Other | 1.14 (0.6 – 2.16) | 0.68 | 1.50 (0.45 – 5.05) | 0.51 | 16.57 (1.89 – 128.13) | 0.009 | 0.91 (0.24 – 3.52) | 0.89 | 0.59 (0.33 - 1.05) | 0.08 |

**Supplementary Table 6** shows the results of multivariable logistic regression from the subcohort analysis (adjusted for grade or stage of training for doctors) as adjusted odds ratios, 95% confidence intervals, and p-values for the association of socio-demographic, health and occupational variables with outcome 1 (redeployment), outcome 2 (received training after redeployment), outcome 3 (received supervision after redeployment), outcome 4 (change in patient contact after redeployment), and outcome 5 (interaction with number of patients with COVID-19).

aORs are adjusted for ethnicity, migration, age, sex, deprivation, and underlying long-term conditions. aOR refers to adjusted Odds Rat**i**

**Supplementary Table 7. Sub-cohort analysis of occupational seniority (for HCWs on the agenda for change pay scale) with redeployment, training, supervision, patient contact, and COVID-19 contact:**

|  | Redeployment: |  | Training: |  | Supervision: |  | Patient Contact: |  | COVID-19 Contact |  |
| --- | --- | --- | --- | --- | --- | --- | --- | --- | --- | --- |
| Agenda for change pay scale | **aOR** | **P value** | **aOR** | **P value** | **aOR** | **P value** | **aOR** | **P**  **value** | **aOR** | **P value** |
| ETHNICITY |  |  |  |  |  |  |  |  |  |  |
| White | Ref | Ref | Ref | Ref | Ref | Ref | Ref | Ref | Ref | ref |
| Black | 0.77 (0.53 – 1.12) | 0.17 | 1.31 (0.61 – 2.83) | 0.49 | 1.26 (0.58 – 2.72) | 0.56 | 0.57 (0.28 – 1.17) | 0.12 | 1.62 (1.15 – 2.27) | 0.006 |
| Asian | 0.83 (0.66 – 1.04) | 0.14 | 0.71 (0.47 – 1.08) | 1.00 | 0.95 (0.61 – 1.47) | 0.81 | 0.93 (0.58 – 1.49) | 0.76 | 1.24 (1.00 – 1.52) | 0.05 |
| Mixed | 1.28 (0.92 – 1.76) | 0.11 | 1.00 (0.56 – 1.80) | 0.11 | 1.17 (0.64 – 2.12) | 0.61 | 0.81 (0.43 – 1.52) | 0.51 | 0.89 (0.63 – 1.21) | 0.42 |
| Other | 1.60 (0.90 – 2.85) | 0.11 | 1.88 (0.60 – 5.84) | 0.28 | 0.93 (0.35 – 2.50) | 0.89 | 1.33 (0.41 – 4.34) | 0.63 | 1.53 (0.83 – 2.81) | 0.17 |
| MIGRATION STATUS |  |  |  |  |  |  |  |  |  |  |
| Born in the UK | Ref | ref | ref | ref | ref | ref | ref | Ref | ref | ref |
| Born abroad | 1.26 (1.06 – 1.49) | 0.01 | 1.23 (0.88 – 1.70) | 0.23 | 1.24 (0.89 – 1.73) | 0.20 | 0.87 (0.61 – 1.24) | 0.43 | 1.15 (0.97 – 1.35) | 0.10 |
| AGE |  |  |  |  |  |  |  |  |  |  |
| 0-31 years | Ref | Ref | ref | ref | ref | ref | ref | ref | ref | ref |
| 31-45 years | 0.92 (0.77 – 1.11) | 0.38 | 0.94 (0.67 – 1.31) | 0.70 | 1.02 (0.73 – 1.44) | 0.90 | 0.52 (0.34 – 0.79) | 0.002 | 0.85 (0.72 – 1.01) | 0.07 |
| 46-60 years | 0.90 (0.75 0 1.09) | 0.30 | 0.99 (0.70 – 1.39) | 0.95 | 0.93 (0.66 – 1.32) | 0.70 | 0.40 (0.26 – 0.62) | <0.001 | 0.64 (0.54 – 0.76) | <0.001 |
| 60 - max | 0.65 (0.47 – 0.89) | 0.008 | 1.28 (0.69 – 2.39) | 0.44 | 1.25 (0.67 – 2.33) | 0.48 | 0.40 (0.20 – 0.80) | 0.01 | 0.47 (0.35 – 0.63) | <0.001 |
| SEX: |  |  |  |  |  |  |  |  |  |  |
| Male | Ref | Ref | Ref | Ref | Ref | Ref | Ref | Ref | Ref | Ref |
| Female | 0.78 (0.66 – 0.92) | 0.004 | 0.93 (0.69 – 1.26) | 0.65 | 0.77 (0.56 – 1.06) | 0.11 | 1.08 (0.77 – 1.52) | 0.65 | 0.71 (0.60 – 0.83) | <0.001 |
| DEPRIVATION: |  |  |  |  |  |  |  |  |  |  |
| 1 (most deprived) | 1.11 (0.88 – 1.41) | 0.39 | 1.14 (0.73 – 1.77) | 0.57 | 1.09 (0.69 – 1.68) | 0.73 | 0.76 (0.46 – 1.26) | 0.30 | 1.12 (0.90 – 1.41) | 0.31 |
| 2 | 1.10 (0.90 – 1.35) | 0.35 | 0.85 (0.57 – 1.26) | 0.42 | 0.90 (0.61 – 1.32) | 0.59 | 0.78 (0.51 – 1.19) | 0.25 | 1.11 (0.92 – 1.36) | 0.28 |
| 3 | Ref | Ref | Ref | Ref | Ref | Ref | Ref | Ref | Ref | Ref |
| 4 | 1.11 (0.89 – 1.33) | 0.39 | 1.01 (0.70 – 1.45) | 0.97 | 1.07 (0.75 – 1.52) | 0.72 | 0.98 (0.65 – 1.47) | 0.92 | 0.93 (0.77 – 1.13) | 0.45 |
| 5 (least deprived) | 1.11(0.90 – 1.31) | 0.36 | 1.09 (0.76 – 1.56) | 0.65 | 0.89 (0.62 – 1.27) | 0.52 | 0.70 (0.47 – 1.03) | 0.07 | 0.87 (0.74 – 1.04) | 0.12 |
| COMORBIDITIES: |  |  |  |  |  |  |  |  |  |  |
| No conditions | Ref | Ref | *(not relevant for training and supervision)* | | | | Ref | Ref | Ref | Ref |
| Long-term conditions that required shielding during COVID-19 | 0.92 (0.79 – 1.06) | 0.26 |  |  |  |  | 0.50 (0.36 – 0.70) | <0.001 | 0.79 (0.67 – 0.93) | 0.004 |
| Other conditions | 0.97 (0.82 – 1.15) | 0.74 |  |  |  |  | 0.91 (0.66 – 1.25) | 0.56 | 0.90 (0.78 – 1.03) | 0.13 |
| Job band |  |  |  |  |  |  |  |  |  |  |
| Band 1-4 | 0.78 (0.58 – 1.05) | 0.10 | 1.06 (0.64 – 1.77) | 0.82 | 0.93 (0.54 – 1.58) | 0.78 | 0.81 (0.43 – 1.47) | 0.49 | 0.69 (0.52 - 0.90) | 0.007 |
| Band 5 | Ref | Ref | Ref | Ref | Ref | Ref | Ref | Ref | Ref | Ref |
| Band 6 | 0.91 (0.76 – 1.08) | 0.26 | 1.11 (0.82 – 1.52) | 0.5 | 0.68 (0.5 – 0.94) | 0.02 | 0.91 (0.63 – 1.32) | 0.62 | 0.62 (0.53 -0.73) | <0.001 |
| Band 7 | 0.86 (0.71 – 1.04) | 0.11 | 0.96 (0.69 – 1.36) | 0.84 | 0.54 (0.38 – 0.76) | <0.001 | 0.71 (0.48 – 1.05) | 0.08 | 0.56 (0.47 - 0.66) | <0.001 |
| Band 8 & 9 | 0.69 (0.55 – 0.87) | 0.001 | 0.83 (0.55 – 1.26) | 0.39 | 0.75 (0.49 – 1.16) | 0.2 | 0.53 (0.33 – 0.83) | 0.006 | 0.32 (0.25 - 0.39) | <0.001 |

**Supplementary Table 7** shows the results of multivariable logistic regression from the subcohort analysis (adjusted for HCWs on agenda for change pay bands) as adjusted odds ratios, 95% confidence intervals, and p-values for the association of socio-demographic, health and occupational variables with outcome 1 (redeployment), outcome 2 (received training after redeployment), outcome 3 (received supervision after redeployment), outcome 4 (change in patient contact after redeployment), and outcome 5 (interaction with number of patients with COVID-19).

aORs are adjusted for ethnicity, migration, age, sex, deprivation, and underlying long-term conditions. aOR refers to adjusted Odds Ratio

**Supplementary Figure 1. Subcohort analysis: The relationship between ethnicity, migration, and doctor’s grade with redeployment after adjustment for demographic and health covariates**

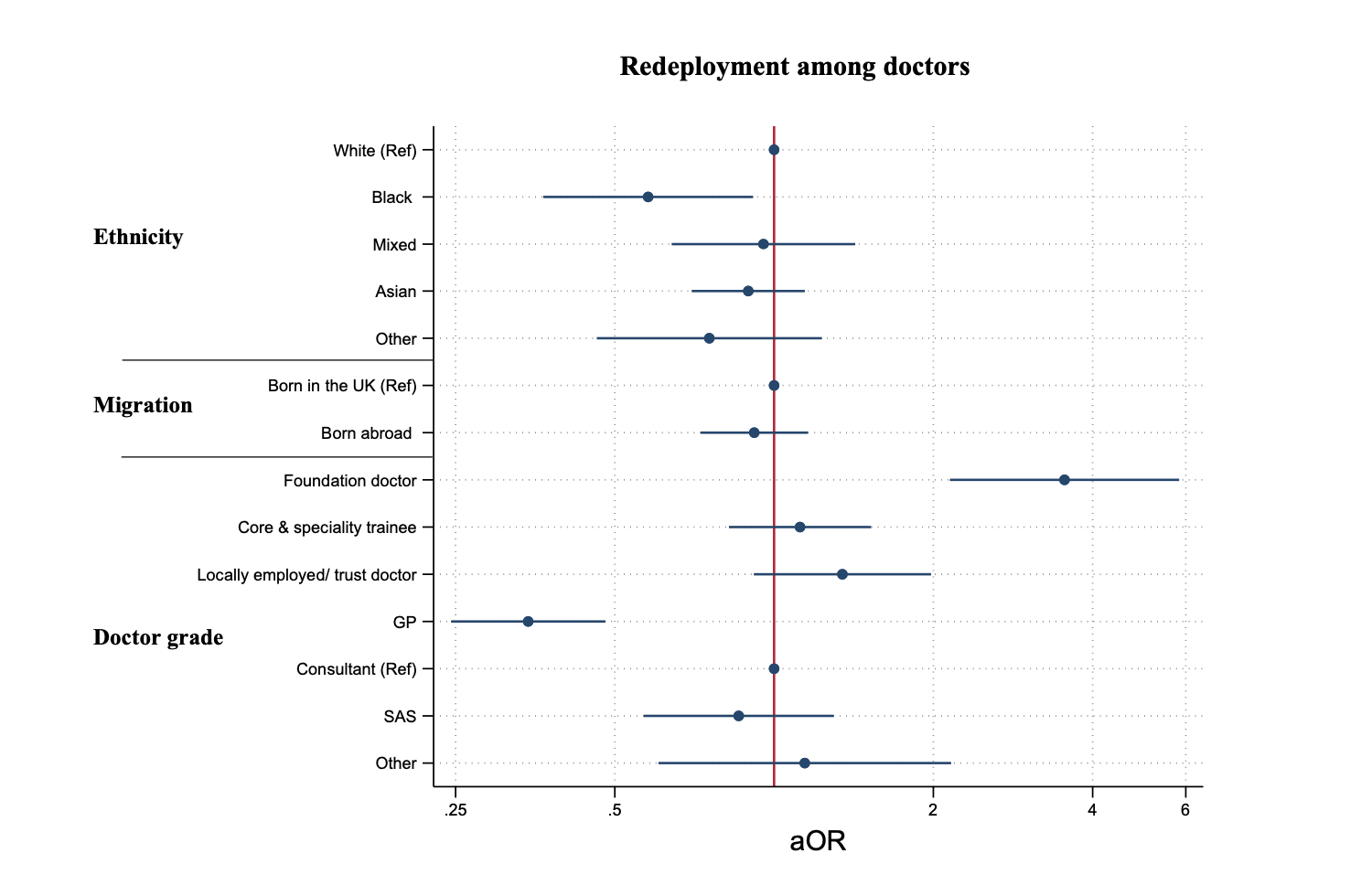

**Supplementary Figure 1** shows adjusted ORs and 95% CI derived from multivariable logistic regression models, to examine the relationship between socio-demographic and occupational (doctor’s grade) variables with redeployment. ORs are adjusted for age, sex, ethnicity, occupation, occupational seniority, migration, deprivation, and underlying long-term health conditions. This figure only shows key results of interest.

**Supplementary Figure 2. Subcohort analysis: The relationship between ethnicity, migration, and job band with redeployment after adjustment for demographic and health covariates**

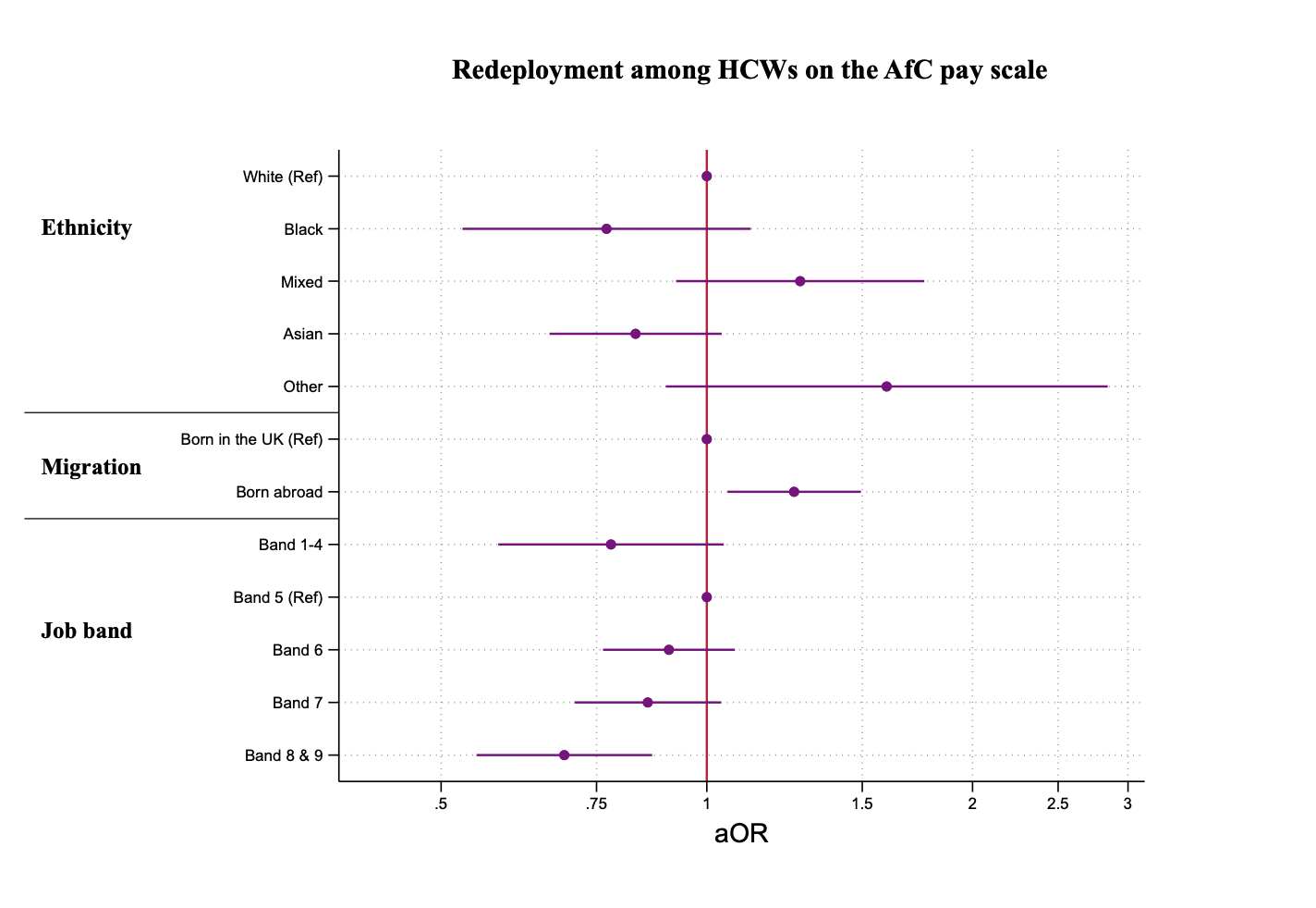

**Supplementary Figure 2** shows adjusted ORs and 95% CI derived from multivariable logistic regression models, to examine the relationship between socio-demographic and occupational (job band) variables with redeployment. ORs are adjusted for age, sex, ethnicity, occupation, occupational seniority, migration, deprivation, and underlying long-term health conditions. This figure only shows key results of interest.

**Supplementary Figure 3. Subcohort analysis: The relationship between ethnicity, migration, and doctor’s grade with training and supervision during redeployment after adjustment for demographic and health covariates**

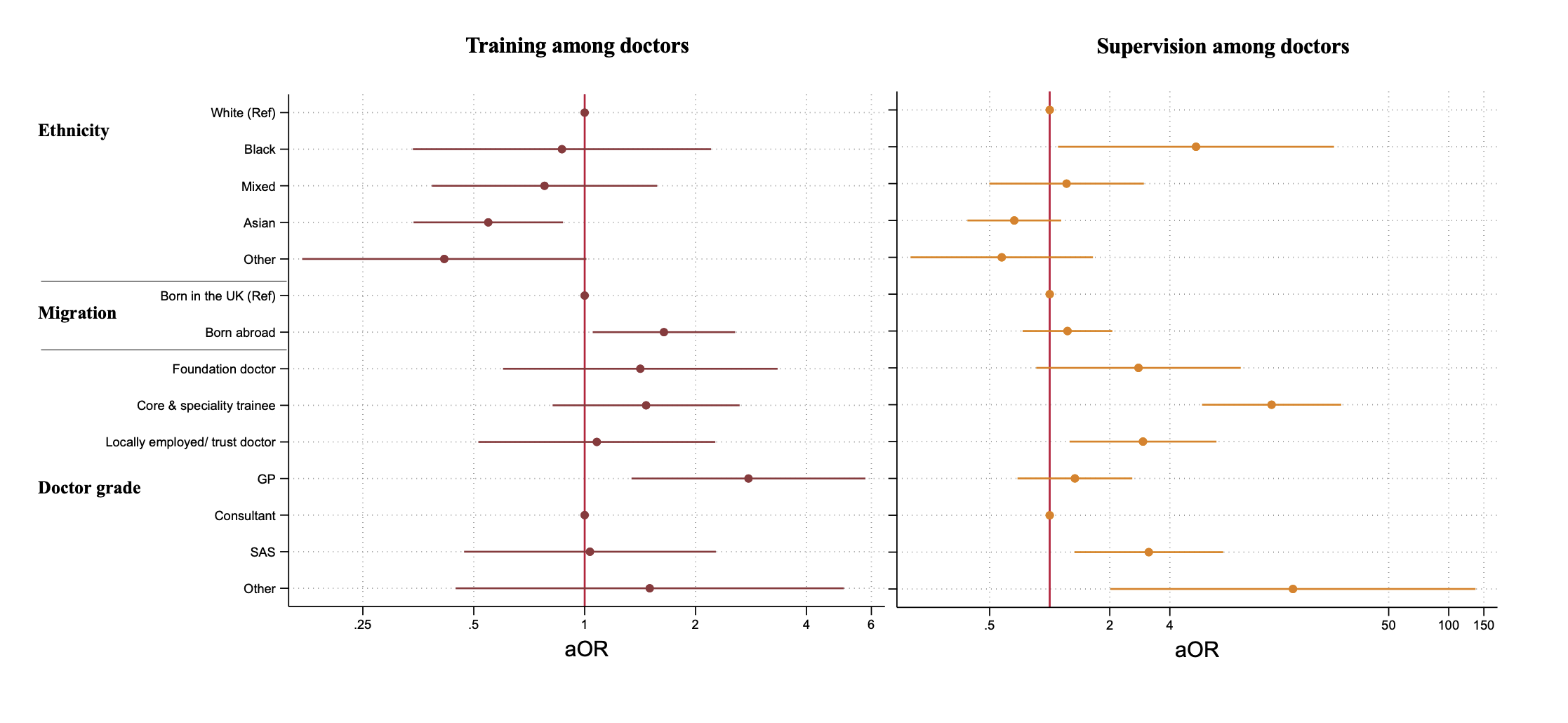

**Supplementary Figure 3** shows adjusted ORs and 95% CI derived from multivariable logistic regression models, to examine the relationship between socio-demographic and occupational (doctor’s grade) variables with training and supervision during. ORs are adjusted for age, sex, ethnicity, occupation, occupational seniority, migration, deprivation, and underlying long-term health conditions. This figure only shows key results of interest.

**Supplementary Figure 4. Subcohort analysis: The relationship between ethnicity, migration, and doctor’s grade with COVID-19 contact after adjustment for demographic and health covariates**

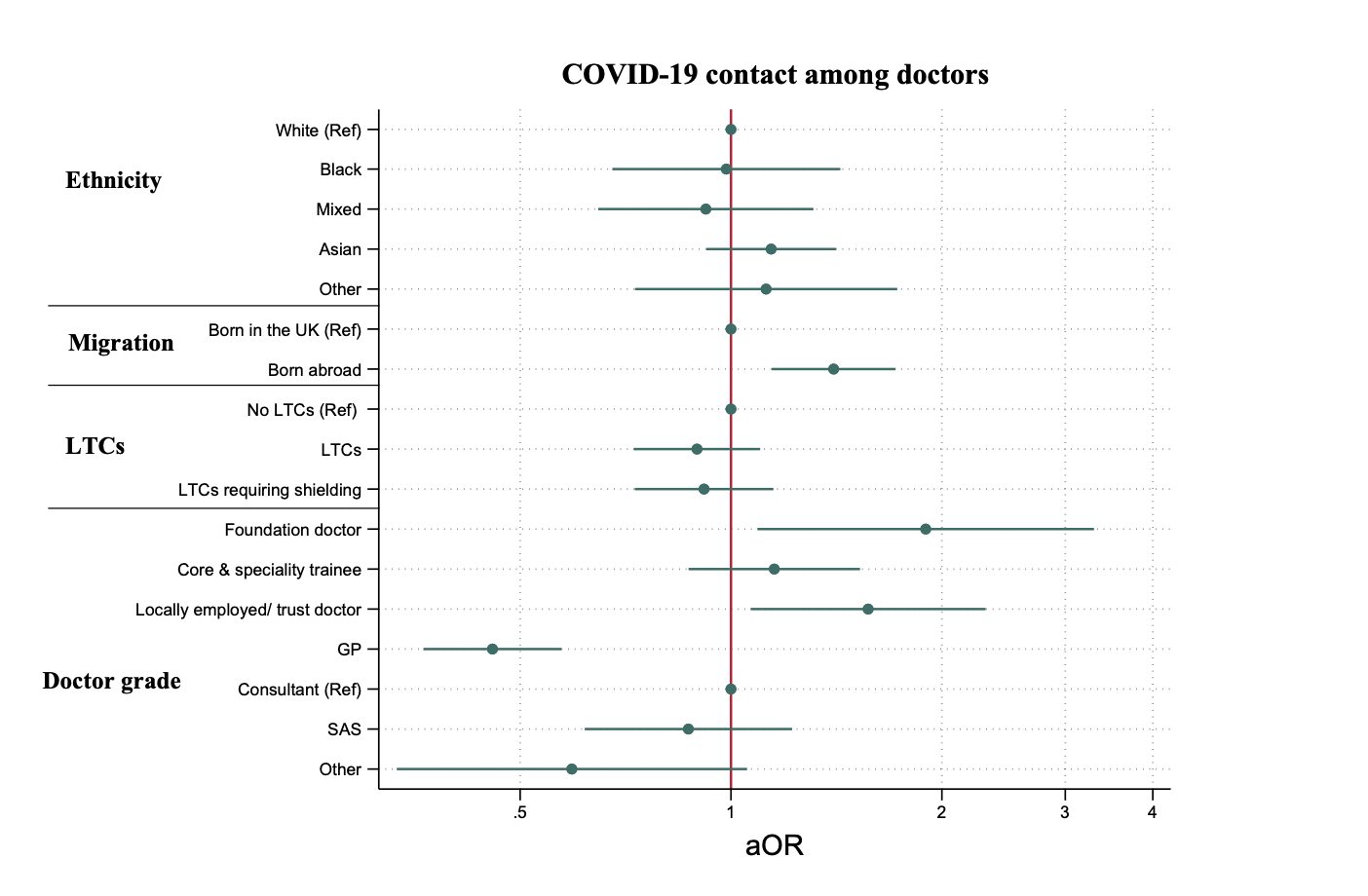

**Supplementary Figure 4** shows adjusted ORs and 95% CI derived from multivariable logistic regression models, to examine the relationship between socio-demographic and occupational (doctor’s grade) variables with COVID-19 contact. ORs are adjusted for age, sex, ethnicity, occupation, occupational seniority, migration, deprivation, and underlying long-term health conditions. This figure only shows key results of interest.

**Supplementary Figure 5. Subcohort analysis: The relationship between ethnicity, migration, and job band with COVID-19 contact after adjustment for demographic and health covariates**

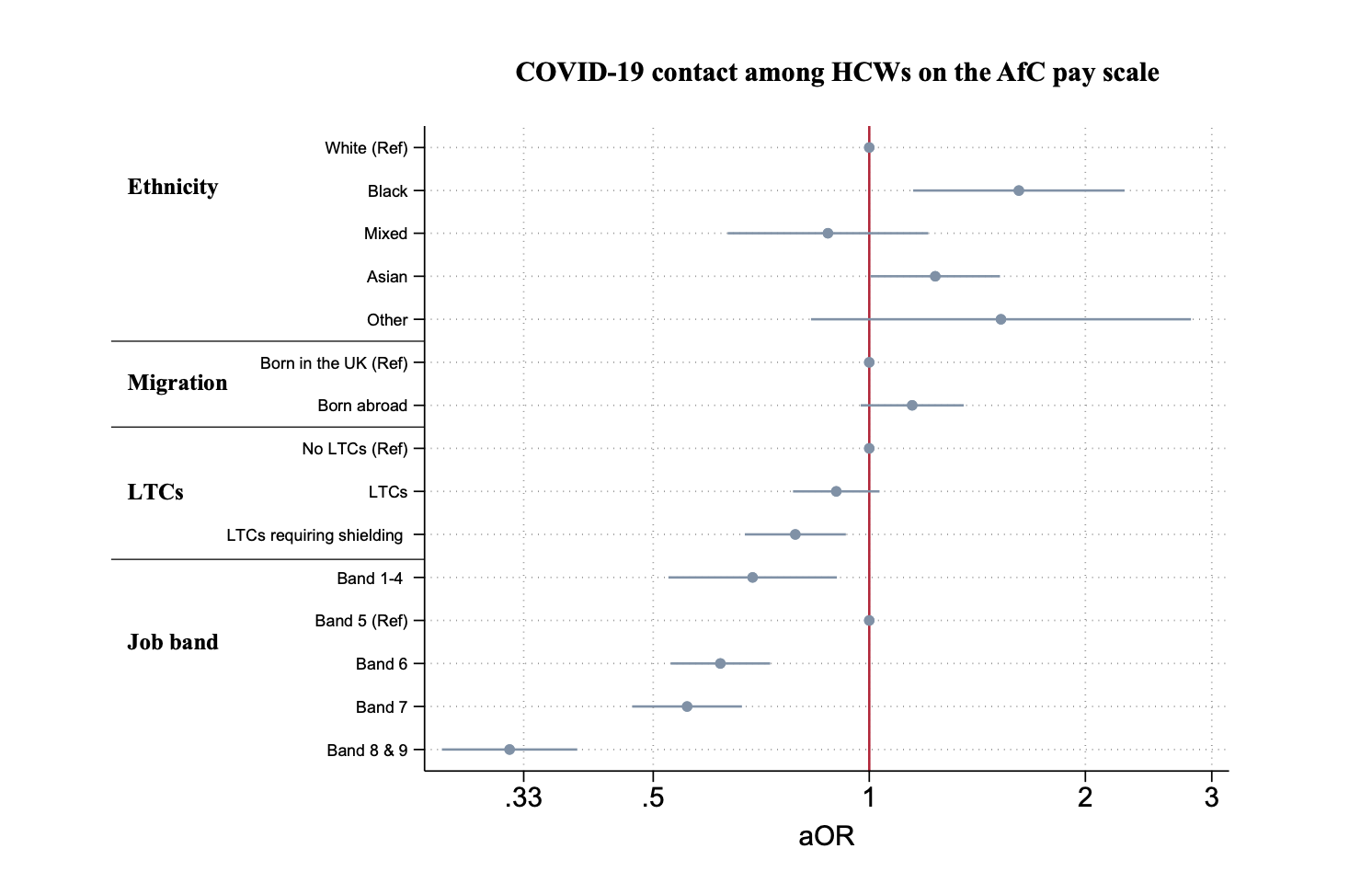

**Supplementary Figure 5** shows adjusted ORs and 95% CI derived from multivariable logistic regression models, to examine the relationship between socio-demographic and occupational (job band) variables with COVID-19 contact. ORs are adjusted for age, sex, ethnicity, occupation, occupational seniority, migration, deprivation, and underlying long-term health conditions. This figure only shows key results of interest.
